# Efficacy of artemisinin-based combination therapy (ACT) in people living with HIV (PLHIV) diagnosed with uncomplicated *Plasmodium* falciparum malaria in Africa: A WWARN systematic review

**DOI:** 10.1101/2025.04.19.25326108

**Authors:** Abena Takyi, Aboubakar Soma, Marianna Przybylska, Eli Harriss, Karen I. Barnes, Prabin Dahal, Philippe J. Guérin, Kasia Stepniewska, Verena I. Carrara

## Abstract

**Background:** Africa bears the highest double burden of HIV and malaria worldwide. In 2023, an estimated 25.9 million people were living with HIV (PLHIV), and 246 million malaria cases were diagnosed in Africa. Malaria patients co-infected with HIV are considered at a higher risk of failing malaria treatment, according to the World Health Organization (WHO) guidelines. This systematic literature review aims to assess the treatment outcomes following artemisinin-based combination therapy (ACT) in PLHIV.

**Methods:** The literature search was conducted up to April 2022 in the following databases: MEDLINE, EMBASE, Web of Science, Cochrane Central, WHO Global Index Medicus, Clinicaltrials.gov, and the WorldWide Antimalarial Resistance Network (WWARN) Clinical Trial Library. Studies describing any malaria treatment outcomes or anti-malarial drug exposure in PLHIV treated for uncomplicated *Plasmodium falciparum* malaria infection were eligible for inclusion.

**Results:** A total of 26 articles describing 19 studies conducted between 2003 and 2017 in six countries were included in this review; it represented 2850 malaria episodes in PLHIV across various transmission settings. The most studied artemisinin-based combination was artemether-lumefantrine (in 16 studies). PLHIV were treated with various antiretroviral therapy (ART) regimens, namely efavirenz (EFV), nevirapine (NVP), atazanavir-ritonavir (ATVr), lopinavir-ritonavir (LPV/r), and/or on prophylaxis with trimethoprim-sulfamethoxazole (TS), or were untreated (in 3 studies). There was no evidence of an increased risk of recrudescence in PLHIV compared to those without HIV. When treated with artemether-lumefantrine, PLHIV receiving LPV/r had a lower risk of malaria recurrence compared to PLHIV on NVP-based or EFV-based ART, or those without HIV. LPV/r increased lumefantrine exposure and EFV-treated patients had a reduced exposure to both artemether and lumefantrine; NVP reduced artemether exposure only.

**Conclusions:** Limited data on ACT outcomes or drug exposure in PLHIV in Africa remains a reality to date, and the effect of antivirals appears inconsistent in the literature. Considering the heterogeneity in study designs, these review findings support conducting an individual patient data meta-analysis to explore the impact of antiretroviral therapy on anti-malarial treatment.

**Trial registration:** The protocol for the original search was published on PROSPERO with registration number CRD42018089860.

## Background

The African continent bears the highest double burden of HIV and malaria worldwide. In 2023, an estimated 25.9 million people lived with HIV (PLHIV) in Africa (65% of all PLHIV) (1), and 246 million malaria cases were diagnosed in the World Health Organization (WHO) African Region, or 93.5% of all cases worldwide (2). A recent publication estimated that in 2020, 1.7 to 2.2 million PLHIV living in 41 African countries may suffer from uncomplicated *Plasmodium falciparum* malaria, contributing to 1.2% of all estimated uncomplicated *P. falciparum*, malaria cases in this region (3).

Naturally acquired immunity against malaria is only partial, and consequently, the immune system of PLHIV residing in malaria-endemic countries has to contend with both HIV and potentially multiple episodes of malaria. As the virus suppresses this acquired partial immunity (4), adult PLHIV may suffer from more frequent symptomatic malaria infections in areas of moderate to high malaria endemicities (5) and are more likely to experience severe disease in low-transmission areas (6). HIV infection has also been shown to increase the risk of malaria infection and has been associated with higher parasite density in pregnancy (7), with a greater risk of malaria re-infection, and treatment failure in adults, even in high transmission areas (5).

Ensuring the efficacy of anti-malarial drugs in a high-risk population, such as PLHIV, is therefore paramount. However, trials specifically studying this coinfection are scarce. A systematic review published in 2011 identified 10 studies that investigated the impact of HIV on anti-malarial treatment response, of which there were only 3 studies that evaluated artemisinin-based combination therapy (ACT) (8). The review found that HIV infection is associated with increased prevalence and severity of clinical malaria and was also associated with impaired response to anti-malarial treatment that was dependent on age, immunosuppression, and previous immunity to malaria. No recent systematic review on efficacy of ACT, the current mainstay of anti-malarial treatment, has been conducted in this population.

Since HIV requires life-long treatment with Highly Active Antiretroviral Therapy (HAART), drug-drug interaction(s) with anti-malarials present possible complications in management of malaria-HIV co-infection. HAART aims to boost the immunity of PLHIV (9). Hence, it is expected that immunity against malaria should also improve. However, the pharmacokinetic properties of anti-malarial drugs may be affected by the presence of HAART, which can alter drug exposure (10, 11). Adverse effects of interactions between ACT and some HAART on liver function and bone marrow suppression have been reported previously (12). Similarly, some drug-drug interactions were previously reported for TS prophylactic treatment, which increases protection against malaria and other opportunistic infections in PLHIV. However, systematic evidence for the effect of drug-drug interactions between ACT and HAART on anti-malarial treatment efficacy is lacking.

The objective of this systematic review was to estimate the efficacy of anti-malarial treatment for uncomplicated *P. falciparum* infection in PLHIV in Africa and compare it with efficacy in HIV-uninfected patients.

## Methods

### Search strategy

The initial search was conducted on 02/09/2019 by a librarian (EH) at the Bodleian Health Care Libraries, University of Oxford, which included all studies published until the search date. The following databases were searched: Ovid MEDLINE, Ovid EMBASE, Web of Science (all Databases), Cochrane Central Register of Controlled Trials, WHO Global Index Medicus and Clinicaltrials.gov. Updates of the search were conducted on 30/10/2020, 01/07/2021, and 28/04/2022 as part of the WWARN Clinical Trial Library (13) and all additional studies published from 01/01/2019 were screened for inclusion. No restrictions were placed on language or publication date. The full list of search terms is available in **Additional File 1** (initial search) and **Additional File 2** (searches in WWARN Clinical Trial Library). Briefly, search terms used in the search strategy included: “Malaria”, “malaria.ti,ab.”, “Plasmodium,” “plasmodium.ti,ab.”, “falciparum,” “Africa,” the name of each African country, or “Central* Africa*” or “West* Africa*” or “East* Africa*” or “North* Africa*” or “South* Africa*” or “sub Saharan Africa*” or “sub-Saharan Africa,*” “artemisinin,” “artemisinin derivative,” the names of each individual component of the ACT, “human immunodeficiency virus infection” or “acquired immune deficiency syndrome” and other related terms.

### Inclusion and exclusion criteria

PLHIV of all ages diagnosed with confirmed uncomplicated *P. falciparum* malaria in Africa were included. Since not every PLHIV has access to life-saving HAART despite its availability for over two decades in sub-Saharan Africa (1), patients on any antiretroviral therapy (ART), TS prophylactic treatment and those not yet on treatment were included. Patients with asymptomatic parasitaemia, severe malaria or unconfirmed malaria were excluded.

The following artemisinin-based combinations were included in this review: artemether-lumefantrine (AL); artesunate-amodiaquine (ASAQ); artesunate-mefloquine (ASMQ); artesunate-sulfadoxine-pyrimethamine (ASSP); dihydroartemisinin-piperaquine (DP); artesunate-pyronaridine (AP).

Studies included were randomized control trials (RCTs), quasi-randomized controlled trials, case-control studies, and longitudinal cohort studies. Pharmacokinetic studies were also included. Animal studies, prevention studies, case reports/case series, retrospective studies, systematic reviews, and literature reviews were excluded.

### Study outcomes and data extraction

The primary outcome was polymerase chain reaction (PCR) adjusted treatment failure (recrudescence) of ACT, as defined by the WHO, on day 28 of treatment for PLHIV (14). Secondary outcomes included other measures of treatment failure such as: PCR confirmed reinfection, recurrence, early treatment failure as well as outcomes recorded on days 42 or 63 of follow-up (14, 15). Pharmacokinetic parameters, if available specifically for PLHIV, were also extracted.

Two reviewers (AT and MP, or AS and MP) independently assessed the eligibility of studies by screening the title and abstract and conducting full text screening of selected studies. Studies were excluded at the title and abstract screening stage if there were no HIV cases, no confirmed uncomplicated *P. falciparum* malaria cases, study sites were not in Africa, prevalence studies, prevention studies, treatment did not include ACT or studies with inappropriate study design. Disagreements between reviewers were resolved by the third reviewer (KS).

Data extracted included study year, site, design, inclusion and exclusion criteria for patients, number of enrolled patients, treatment regimens, number of patients treated with each regimen, and reported outcome. For each reported outcome and each arm/patient subgroup, number of patients evaluated, day of assessment and treatment efficacy results were extracted. When provided, measurements of treatment differences in anti-malarial treatment efficacy, such as Hazard Ratios (HR), Risk Ratio (RR) or Odds Ratio (OR) with 95% confidence intervals (CI), and p-value were extracted. Any estimates of pharmacokinetics (PK) parameters for anti-malarial drugs from pharmacokinetic studies were included.

### Risk of bias assessment

Risk of bias in individual studies was assessed using the Cochrane tools RoB 2 for randomized studies (16) and ROBINS-I for non-randomized studies (17). A set of signalling questions was used to make a judgement on the likely extent of bias for each of the studies across different domains under consideration (see **Additional Files 3 and 4** for a full set of signalling questions and judgments for each of the studies). Certainty of evidence for each outcome was assessed according to the GRADE guidelines (18).

### Statistical analysis

Due to the small number of studies and limited information provided in publications, only descriptive analysis was carried out for the majority of outcomes. Meta-analysis could only be conducted to compare lumefantrine concentrations on day 7 between different ART regimens and to compare artemether, or its metabolite dihydroartemisinin, exposure between PLHIV treated with NVP and HIV-uninfected patients. Fixed effect models using the method of Mantel and Haenszel were fitted and I-squared was used as a relative measure of heterogeneity between studies.

Since studies included in this review were conducted in high or moderate transmission intensity areas, efficacy estimates were presented for recrudescence only, while relative estimates (OR, HR) were presented for any outcomes for comparison between PLHIV and HIV-uninfected patients, or different ART regimens in PLHIV. Where raw data were available, the calculations were conducted to estimate proportion (95% CI) of patients with the outcome of interest; OR (95% CI) were calculated from proportions, and HR were calculated from the Kaplan-Meier (KM) estimates as outlined in Klein *et al.* (19). Interquartile ranges (IQR) of the drug levels, if not provided, were estimated from other reported parameters such as range (after logarithmic transformation, using a method outlined in Hozo *et al*. (20) and implemented in an online calculator (21)), or from reported mean and its 95% CI (assuming normal distribution). Similarly, mean and 95% CI were estimated from median and IQR as proposed by Wan *et al.* (22).

## Results

A total of 9,950 articles were identified for screening, 990 were included in the full text screening (**Fig. 1**). Twenty-six articles (originating from 19 studies) were identified for inclusion in the review. These studies were conducted between 2003 and 2017 in various endemicity areas in Uganda (n=10), Nigeria (n=4), Zambia (n=2) and Tanzania (n=1). There were two multi-country studies (one in Malawi and Uganda, and the other in Malawi and Mozambique) (**Table 1**).

**Fig. 1.**
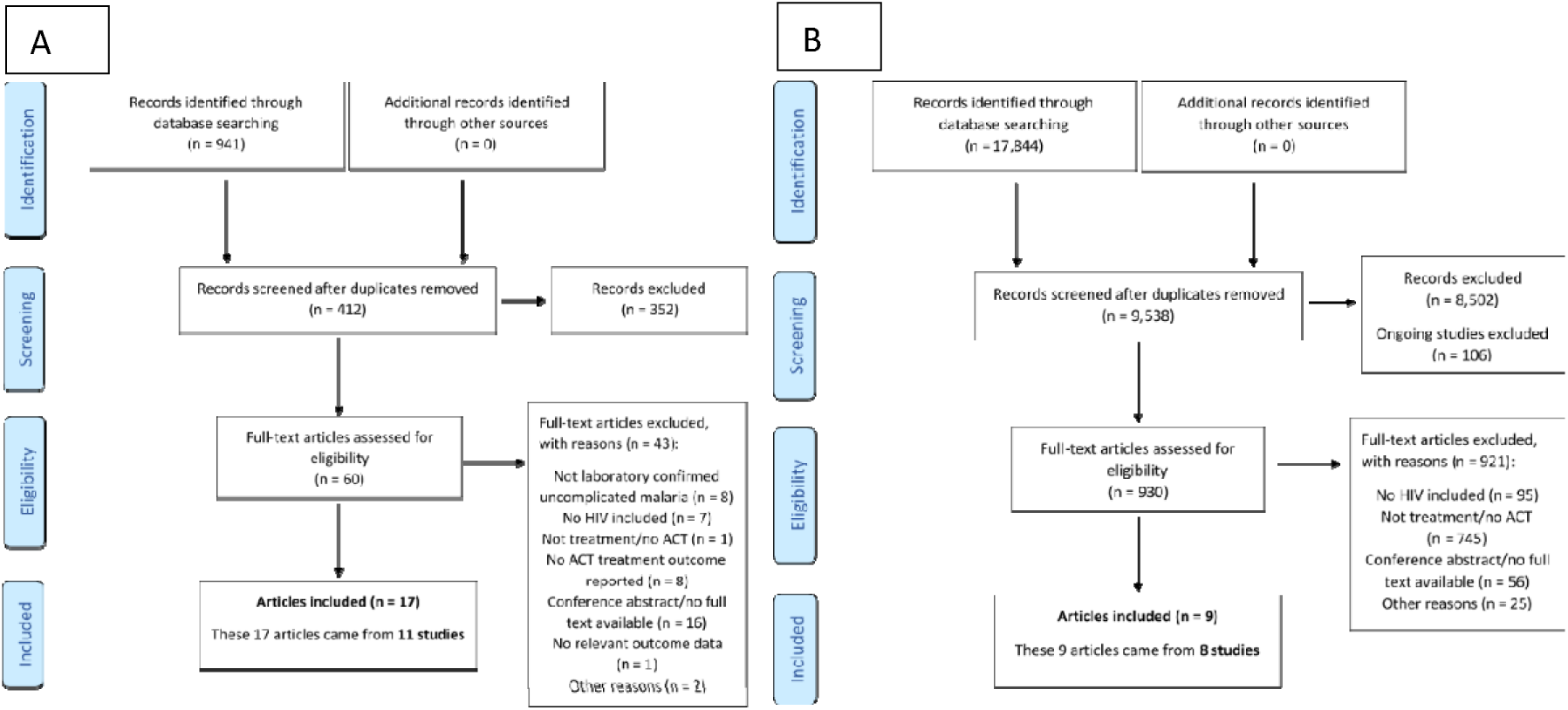
PRISMA profile for systematic review. On the left (panel A), initial search on MEDLINE, EMBASE, Web of Science (all Databases), Cochrane Central, WHO Global Index Medicus and Clinicaltrials.gov. On the right (panel B), subsequent searches performed in the WWARN Clinical Trial Library.

**Table 1.** Summary of the 26 articles included in the review and their associated study (Study ID)

| Study ID | First author, date and PMID of all included articles | Study years | Country | Transmission intensity | Study design | Patients enrolled | Age (years) | Pregnancy included | HIV-uninfected Included | Malaria drug | HIV drug | Total malaria episodes in PLHIV | Total malaria episodes overall | Clinical study | PCR | PK parameters |
| --- | --- | --- | --- | --- | --- | --- | --- | --- | --- | --- | --- | --- | --- | --- | --- | --- |
| 1 | Huang-2017; 29065172 (40) | 2012-2014 | Malawi<br>Uganda | Not provided | Observational, non-randomized study | 39 | 3.6-12 | N/A | Yes (historical controls) | AL | NVP | 15 | 35 | No | N/A | Artemether DHA LUM |
| 2 | 1. Maganda-2014; 24885714 (25)<br>2. Maganda-2015; 25906774 (38)<br>3. Maganda-2016; 25963334 (58) | 2010-2012 | Tanzania | Moderate | Open-label, parallel three-arm study | 269 | 19-67 | No | No | AL | EFV<br>NVP<br>None | 269 | 269 | Yes | No | LUM |
| 3 | Gasarira-2008; 18444813 (45) | 2005-2006 | Uganda | Moderate | Longitudinal, 2 prospective cohorts | 293 | 1-10 | N/A | Yes | AS<br>AQ | TS+ART | 35 | 293 | Yes | Yes | No |
| 4 | Sambol-2015; 25732044 (39) | 2008 | Uganda | High | Part of longitudinal, open-label RCT | 107 | 0.5-2 | N/A | Yes | DP | TS+ART | 12<sup>3</sup> | 218 | No | N/A | PPQ |
| 5 | 1. Arinaitwe-2009; 19877969 (59)<br>2. Verret-2011; 21383095 (29) | 2007-2009 | Uganda | High | Longitudinal, open-label RCT | 292 | 0.33-3 | N/A | Yes | AL<br>DP | TS<br>TS+ART | 172 | 2,013 | Yes | Yes<sub>1</sub> | No |
| 6 | 1. Kakuru-2013; 23382157 (36)<br>2. Wanzira-2014; 24825870 (30) | 2007-2011 | Uganda | High | Longitudinal, open-label RCT | 312 | 0.33-5 | N/A | Yes | AL<br>DP | TS+ART | 44<sup>4</sup> | 5,564 | Yes | No | No |
| 7 | Muhindo-2014; 24468007 (35) | 2011-2012 | Uganda | High | Longitudinal, open-label RCT | 202 | 4-5 | N/A | Yes | AL<br>DP | TS+ART | 38 | 770 | Yes | No | No |
| 8 | Kakuru-2014; 24759826 | 2009- | Uganda | High | Longitudinal, 2 open- | 166 | 0.12-6 | N/A | No | AL | EFV | 938 | 938 | Yes | No | No |
|  | (31) | 2013<br>2007-2012 |  |  | label RCTs |  |  |  |  | DP | NVP<br>LPV/r<br>None |  |  |  |  |  |
| 9 | Achan-2012; 23190222<br>(33) | 2009-2011 | Uganda | High | Longitudinal, open-label<br>RCT | 170 | 0.5-7 | N/A | No | AL | EFV<br>NVP<br>LPV/r | 281 | 281 | Yes | No | LUM |
| 10 | Musoke-2012; ISSN:<br>1816-3319 (37) | 2010-2011 | Uganda | High | Open-label, parallel<br>three-arm study | 36 | Adults | No | No | AL | TS<br>TS+EFV | 36 | 36 | No | N/A | LUM |
| 11 | 1. Kajubi-2016;<br>28018925 (34)<br>2. Parikh-2016;<br>27143666 (26) | 2011-2014 | Uganda | High | Longitudinal,<br>prospective PK/PD<br>study | 252 | 1.1-8.6 | N/A | Yes | AL | EFV<br>NVP<br>LPV/r | 118 | 134 | Yes | Yes | Artemether<br>DHA<br>LUM |
| 12 | 1. Van Geertruyden-2006; 16960779 (24)<br>2. Van Geertruyden-2009; 19922664 (60) | 2003-2005 | Zambia | Moderate | RCT | 970 | Adults | No | Yes | AL<br>SP | None | 320 | 970 | Yes | Yes | No |
| 13 | Usman-2021; 33755736<br>(61) | Not<br>provided | Nigeria | Not<br>provided | PK parallel study | 20 | Adults<br>≥18 | No | Yes | AL | ATV/r | 10 | 20 | No | N/A | LUM |
| 14 | 1. Adegbola-2018;<br>30082286 (62)<br>2. Adegbola-2020;<br>32209837 (63) | 2016-2017 | Nigeria | Unstable | PK parallel study | 69 | 18-45 | Yes | No | AL | EFV | 52 | 52 | No | N/A | LUM<br>Desbutyl-Lum |
| 15 | Hugues-2020; 31929402<br>(28) | 2012-2014 | Uganda | High | PK parallel study | 39 | ≥16 | Yes,<br>all | Yes | AL | EFV | 39 | 39 | Yes | No | Artemether<br>DHA<br>LUM |
| 16 | Usman-2020; 32921396<br>(54) | Not<br>provided | Nigeria | Not<br>provided | PK parallel study | 40 | ≥18 | No | Yes | AL | EFV<br>NVP<br>LPV/r | 30 | 40 | No | N/A | LUM |
| 17 | Banda-2019; 31126288 | 2014- | Zambia | Moderate | Single-arm | 152 | 15-65 | No | No | AL | EFV | 152 | 152 | Yes | Yes | No |
|  | (64) | 2015 |  | te-High | interventional study |  |  |  |  |  |  |  |  |  |  |  |
| 18 | Sevene-2019; 31429785 (32) | 2013-2015 | Malawi Mozambique | Moderate-High | Single-arm interventional study | 221 | 15-65 | No | No | DP | EFV NVP | 221 | 221 | Yes | Yes | No |
| 19 | Chijioke-Nwauche-2013; 23774430 (27) <sup>2</sup> | 2010-2011 | Nigeria | Holoendemic | PK parallel study | 167 | 16-65 | Not provided | Yes | AL | EFV NVP | 68 | 167 | Yes | No | LUM |

Study 8 combines a set of patients from Study 4 and an extension of Study 8 is reported in Study 9
Studies will be referred to in the text (i.e. ID 1) and the relevant article afferent to the study will be referenced
<sup>1</sup> PCR done, but no PCR corrected results presented
<sup>2</sup> All malaria slides positive by microscopy, but 78.1% were aparasitaemic by nested-PCR
<sup>3</sup> 12 children were HIV-infected and had at least one malaria episode; total number of episodes not reported
<sup>4</sup> 44 children were HIV-infected and had at least one malaria episode; total number of episodes not reported
ACT: artemisinin combination therapy; AL: artemether-lumefantrine; ART: antiretroviral therapy; ASAQ: artesunate-amodiaquine; ATV/r: atazanavir-ritonavir; Desbutyl-Lum: desbutyl-lumefantrine; DHA: dihydroartemisinin; DP: dihydroartemisinin-piperaquine; EFV: efavirenz; LUM: lumefantrine; LPV/r: lopinavir/ritonavir; N/A: information not available; NVP: nevirapine; PLHIV: people living with HIV; PK: pharmacokinetics parameters; PCR: polymerase chain reaction (adjusted treatment failure); PPQ: piperaquine; SP: sulfadoxine-pyrimethamine; TS: trimethoprim-sulfamethoxazole prophylactic treatment

The artemisinin-based combinations studied included AL in 16 studies (including 10 pharmacokinetic studies of lumefantrine, and of artemether and its metabolite dihydroartemisinin in 3 of them), DP in six studies (5 clinical and one study reporting piperaquine pharmacokinetics) and ASAQ in one clinical study. In total, 12,450 malaria episodes were reported in the included studies, with 79% of the episodes (n=9,784/12,450) reported in longitudinal cohorts of children studied in Tororo, Uganda (study ID 4-9, **Table 1**). There were 2,850 malaria episodes among PLHIV.

Seven studies (5 in adults and 2 in children) included only PLHIV; two studies included pregnant women. Eight studies compared malaria outcomes under different ART regimens, namely efavirenz (EFV), nevirapine (NVP), lopinavir-ritonavir (LPV/r) or atazanavir-ritonavir (ATV/r), and/or prophylaxis with TS. Detailed description of HIV-related inclusion criteria is available in **Additional File 5**. Most studies did not have restrictions on the CD4 count in PLHIV at enrolment, and only seven studies provided a baseline CD4 count: in those, PLHIV were on ART for at least two weeks prior to enrolment with a reported median CD4 count >200 cells/mm^3^ or a CD4 percentage >20% (23).

### Late treatment failure

Eleven studies presented findings regarding late treatment failure. Risk of recurrence was compared between malaria patients with and without HIV infection in seven studies (**Table 2**).

**Table 2.**
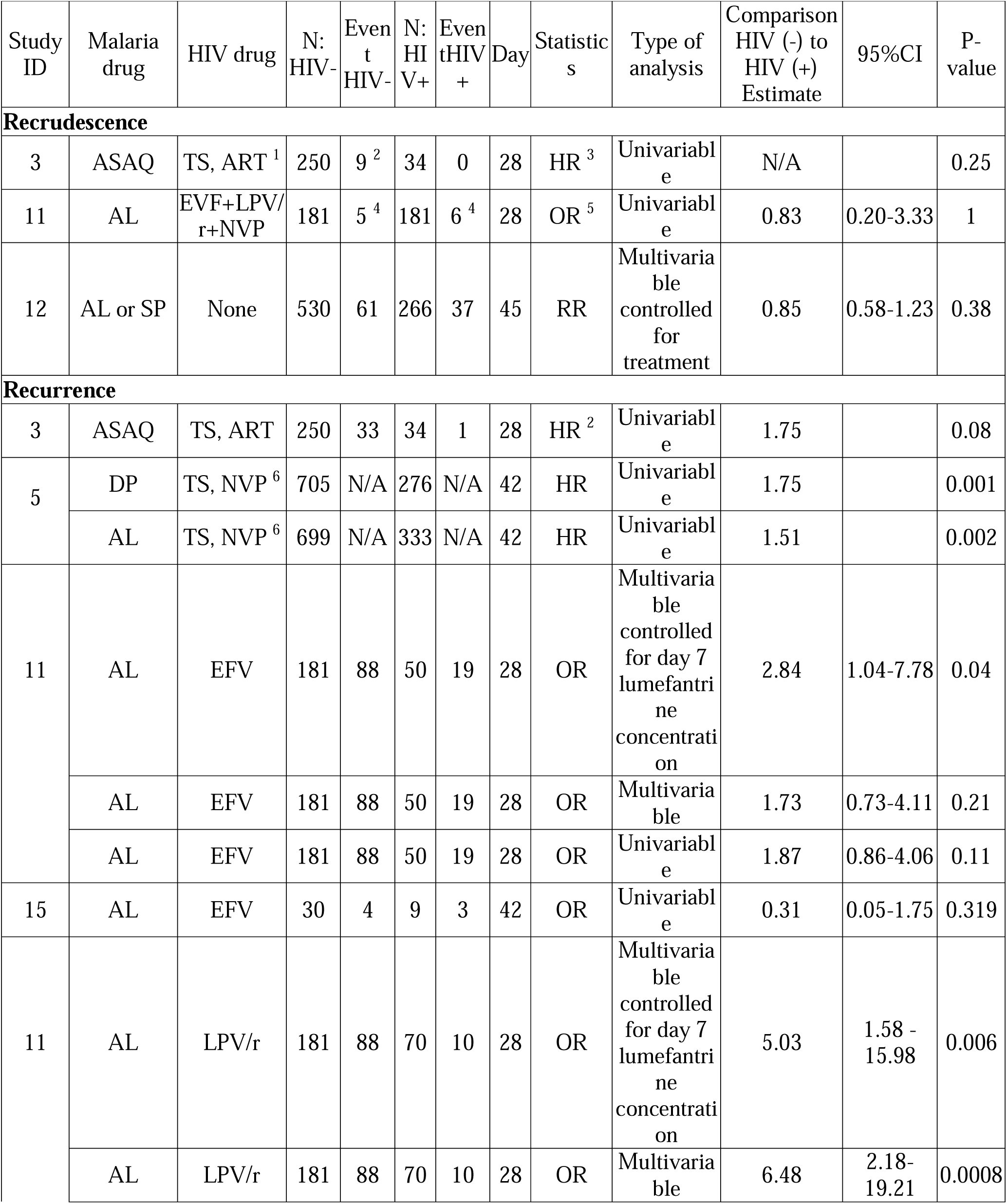
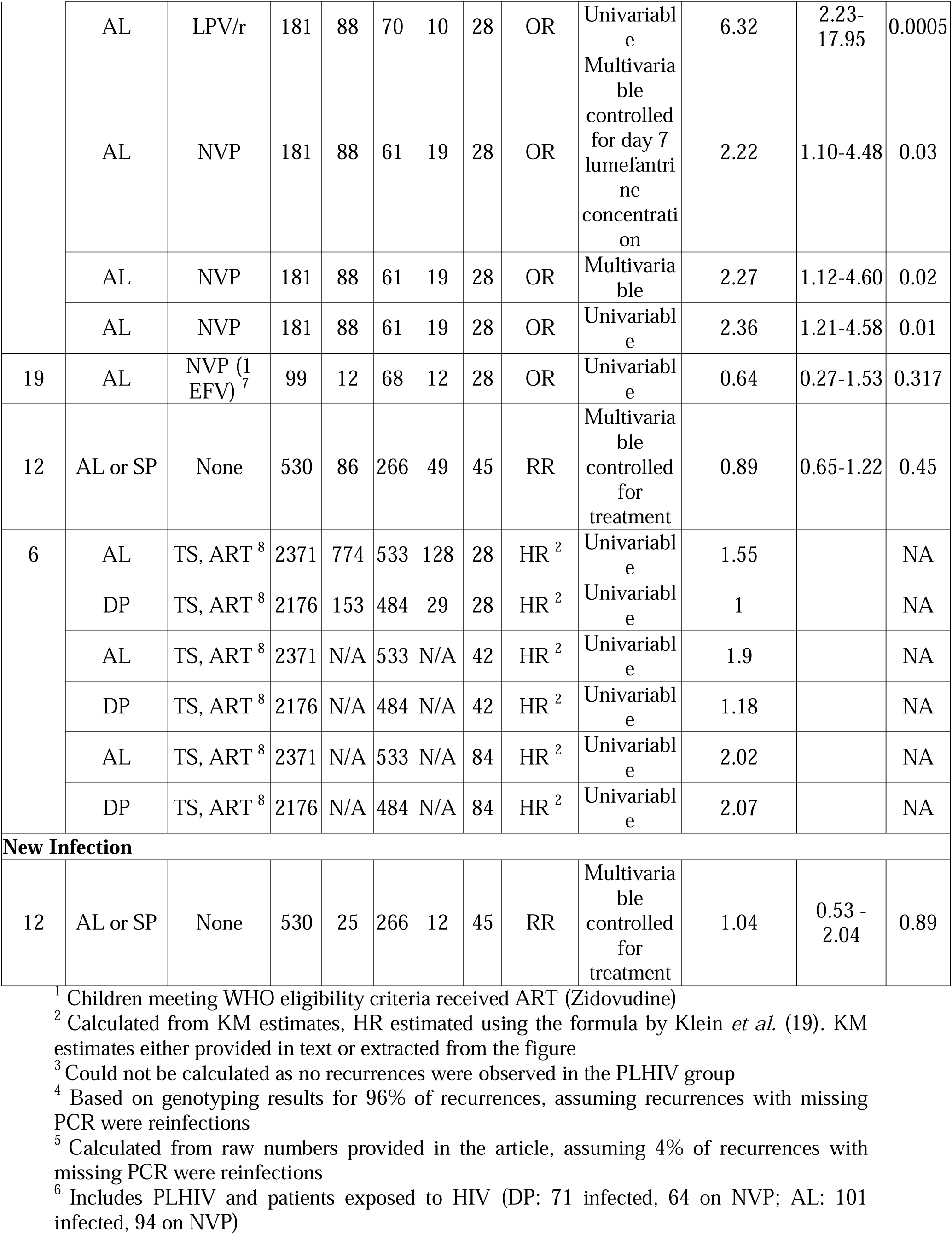
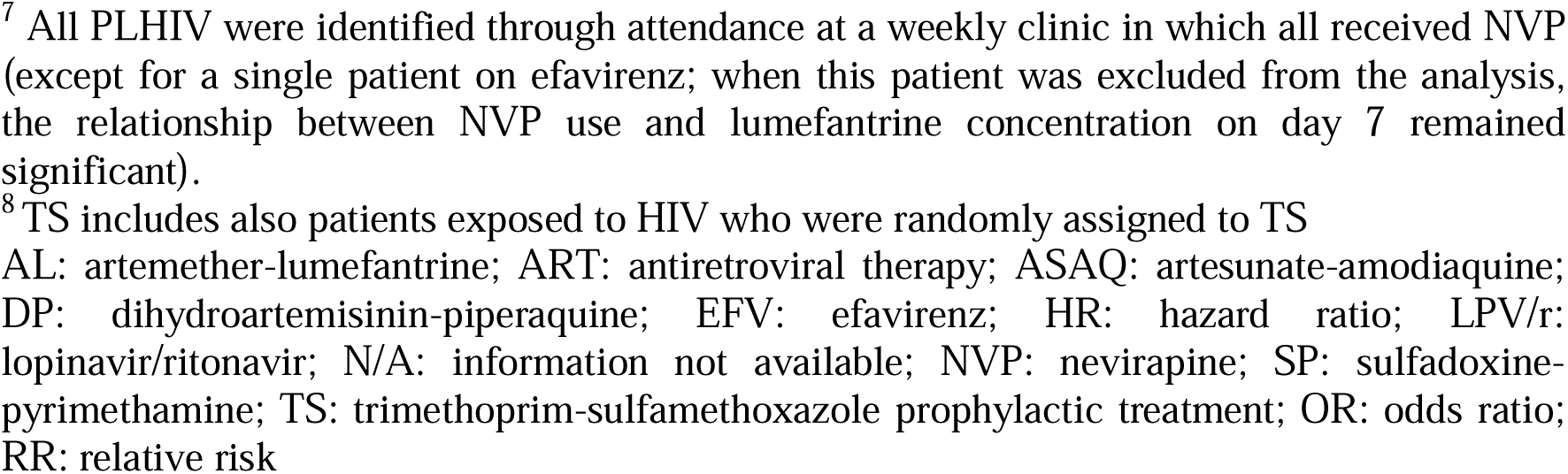
Comparison of anti-malarial treatment efficacy between PLHIV and HIV-uninfected malaria patients.

One study (ID 12) conducted in Zambian adults investigated malaria efficacy in PLHIV not yet on ART. This study treated malaria with AL or SP, and presented results for the combined treatment arms on day 45 (24); after adjusting for treatment, there were no significant differences between PLHIV and HIV-uninfected patients with respect to risk of recurrence, recrudescence or reinfection. However, among PLHIV, the risk of malaria treatment failure (unadjusted for recrudescence or reinfection) on day 45 was found to be 2.24-fold higher among those with CD4 cell count <300 cells/µL compared to those with CD4 cell count ≥300 cells/µL (RR 2.24, 95%CI 1.20 – 4.17, p-value=0.01). This finding was not confirmed in the other study (ID 2) as no significant difference in risk of recurrence at day 28 was observed between patients with CD4 cell count above or below 350 cells/µL (25). Our search identified only one study from Uganda conducted in children (ID 11), which explored the effect of HIV and different ART treatments on malaria treatment outcomes. This study found the risk of recurrence in children living with HIV on ART was significantly lower than in children who were HIV-uninfected (26). The overall odds ratio (OR) of recurrence (adjusted for day 7 lumefantrine concentration) among the HIV-uninfected group was higher compared to PLHIV on LPV/r, NVP or EFV-based regimens, and these were 5.03 (1.58-15.98, p=0.006), 2.22 (1.10-4.48, p=0.003) and 2.84 (1.04 −7.78, p=0.04) respectively (26). When recurrence was compared among the 3 ART regimens, children on EFV had an adjusted OR (AOR) 3.74 times that of LPV/r. A similar recurrence was observed between LPV/r and NVP. In children treated with EFV, the observed frequency of recurrence was similar to the HIV-uninfected group and this was due to lower lumefantrine drug exposure. The comparisons between all treatment groups were not significantly different for recrudescence. Two further studies in adults treated with AL found a higher proportion of recurrence in PLHIV receiving EFV (ID 15 in Uganda) or NVP (ID 19 in Nigeria), compared to HIV-uninfected patients; however, the differences were not significant (27, 28).

Three other studies (ID 3, ID 5, ID 6) compared outcomes in PLHIV on TS prophylaxis (some of whom were also on ART as per country protocols) to HIV-uninfected patients. These three studies showed consistent results of higher risk of recurrence in HIV-uninfected patients with corresponding HR on day 28 of 1.5 for AL and 1.75 for ASAQ, with a pooled OR of 1.35 (95% CI 1.16-1.56, p<0.001, I^2^=0.0%, chi-square test for heterogeneity=0.376), based on available observed proportions (ID 3, ID 6, AL or ASAQ) and ignoring losses to follow-up (as no information was available). For patients treated with DP, no difference (HR = 1.75) was observed until day 42 in the study by Verret *et al.* (29), and an estimated HR of 1.2 (derived from KM curves) was reported in the study by Wanzira *et al.* (30). The HR reached 2 by day 84 in the study by Wanzira *et al.* (30). Confidence intervals for HR were not provided or could not be calculated, and KM estimates could not be pooled as the standard errors were not available.

**Table 3** presents the reported estimates of PCR-corrected risk of recrudescence in PLHIV in five studies. Estimates for recurrence are not provided as they are driven by the malaria transmission intensity which is moderate to high in all study sites (**Additional File 6**).

**Table 3.** Estimated PCR-corrected risk of recrudescence in PLHIV co-infected with malaria.

| Study ID | Day | Malaria drug | HIV drug | PLHIV group |  | % | 95%CI | HIV-uninfected comparator |  | % | 95%CI |
| --- | --- | --- | --- | --- | --- | --- | --- | --- | --- | --- | --- |
|  |  |  |  | N | n |  |  | N | n |  |  |
| 3 | 28 | ASAQ | TS+ART | 35 | 0 | 0.0 | 0-10.0 | 258 |  | 3.6<sup>1</sup> | 1.9-6.9<sub>1</sub> |
| 11 | 28 | AL | NVP | 61 | 3 | 4.9 | 1.0-13.7 | 181 | 5 | 2.8 | 1.2-6.3 |
|  | 28 | AL | EFV | 50 | 1 | 2.0 | 0.1-10.6 | 181 | 5 | 2.8 | 1.2-6.3 |
|  | 28 | AL | LPV/r | 70 | 2 | 2.9 | 0.3-9.9 | 181 | 5 | 2.8 | 1.2-6.3 |
| 12 | 45 | AL or SP | None | 266 | 37 | 13.9 | 10.0-18.7 | 530 | 61 | 11.5 | 8.9-14.5 |
| 17 | 42 | AL | EFV | 134<sup>2</sup> | 0 | 0.0 | 0.0-2.8 |  |  |  |  |
| 18 | 42 | DP | EFV | 158<sup>3</sup> | 0 | 0.0 | 0.0-2.4 |  |  |  |  |
|  | 42 | DP | NVP | 61 | 0 | 0.0 | 0.0-6.9 |  |  |  |  |

N=number of patients enrolled, and n=number of failures. If only n/N were provided in the publication, proportion and 95% CI for proportion were calculated (Wilson method) assuming no losses to follow-up
<sup>1</sup> KM estimates are provided as reported in the publication
<sup>2</sup> Excludes 10 lost to follow-up and 8 with PCR results indeterminate or unavailable
<sup>3</sup> Excludes one early treatment failure and one with PCR result not available
AL: artemether-lumefantrine; ART: antiretroviral therapy; ASAQ: artesunate-amodiaquine; DP: dihydroartemisinin-piperaquine; EFV: efavirenz; LPV/r: lopinavir/ritonavir; NVP: nevirapine; SP: sulfadoxine-pyrimethamine; TS: trimethoprim-sulfamethoxazole prophylactic treatment

Five studies (ID 2, ID 8, ID 9, ID 11, ID 18) compared the risk of recurrence after ACT in PLHIV on different ART regimens (**Table 4**). A slightly higher proportion of recurrent malaria was reported in patients on EFV compared to patients on NVP by day 28 (26, 31) or day 42 (32); however, none of the comparisons were statistically significant. Compared to LPV/r, NVP and EFV had approximately 3-fold higher risk of recurrence by day 28 (26, 31, 33); this finding was no longer apparent once the comparison was adjusted for lumefantrine concentration on day 7 in study ID 11 (26). Study ID 9 (33) showed that lower concentrations of lumefantrine on day 7 were observed in EFV or NVP treatment groups compared to LPV/r (further discussed in the pharmacokinetic section). Meta-analysis was not attempted as raw data was either not available or different effect measures (OR, HR) were reported.

**Table 4.** Comparison of risk of recurrence of parasitaemia in PLHIV on different antiretroviral therapies or not yet on treatment.

| Study ID | ACT | ART Group 1 | ART Group 2 | N1 | Events1 | N2 | Events2 | Day | Statistics | Type of analysis | Estimate | 95%CI | P-value |
| --- | --- | --- | --- | --- | --- | --- | --- | --- | --- | --- | --- | --- | --- |
| 9 | AL | NVP or EFV | LPV/r | 86 | N/A | 84 | N/A | 63 | HR | Univariable | 2.44 | 1.32-4.54 | 0.004 |
|  | AL | NVP or EFV | LPV/r | 86 | N/A | 84 | N/A | 28 | HR | Univariable | 3.23 | 1.47-7.14 | 0.004 |
| 2 | AL | NVP | None | 125 | 3 | 73 | 4 | 28 | RR | Univariable | 0.4 | 0.29-0.9 | 0.53 |
|  | AL | EFV | None | 63 | 11 | 73 | 4 | 28 | RR | Univariable | 3.2 | 2.4-7.8 | <0.001 |
|  | AL | EFV | None | 63 | 11 | 73 | 4 | 28 | HR | Univariable | 19.11 | 10.5-34.5 | <0.01 |
|  | AL | NVP | None | 125 | 3 | 73 | 4 | 28 | HR | Univariable | 2.44 | 0.79-7.6 | 0.53 |
| 8 | AL | EFV | NVP | 102 | N/A | 410 | N/A | 28 | HR | Multivariable controlled for age | 1.76 | 0.99-3.13 | 0.06 |
|  | AL | NVP | LPV/r | 410 | N/A | 249 | N/A | 28 | HR | Multivariable controlled for age | 2.65 | 1.27-5.26 | 0.009 |
|  | AL or DP | NVP | None | 567 | N/A | 20 | N/A | 28 | HR | Multivariate controlled for age | 6.25 | 0.83-50.0 | 0.08 |
| 11 | AL | EFV | LPV/r | 50 | 19 | 70 | 10 | 28 | OR | Univariable | 3.38 | 1.01-11.35 | 0.49 |
|  | AL | EFV | LPV/r | 50 | 19 | 70 | 10 | 28 | OR | Multivariable | 3.74 | 1.02-13.74 | 0.04 |
|  | AL | EFV | LPV/r | 50 | 19 | 70 | 10 | 28 | OR | Multivariable controlled for day 7 lumefantrine concentration | 1.77 | 0.36-8.81 | 0.48 |
|  | AL | NVP | LPV/r | 61 | 19 | 70 | 10 | 28 | OR | Univariable | 2.68 | 0.84-8.56 | 0.1 |
|  | AL | NVP | LPV/r | 61 | 19 | 70 | 10 | 28 | OR | Multivariable | 2.86 | 0.85-9.60 | 0.09 |
|  | AL | NVP | LPV/r | 61 | 19 | 70 | 10 | 28 | OR | Multivariable controlled for day 7 lumefantrine concentration | 2.26 | 0.64-8.05 | 0.21 |
|  | AL | EFV | NVP | 50 | 19 | 61 | 19 | 28 | OR | Univariable | 1.26 | 0.5-3.17 | 0.62 |
|  | AL | EFV | NVP | 50 | 19 | 61 | 19 | 28 | OR | Multivariable | 1.31 | 0.49-3.51 | 0.59 |
|  | AL | EFV | NVP | 50 | 19 | 61 | 19 | 28 | OR | Multivariable controlled for day 7 | 0.78 | 0.32-2.37 | 0.66 |
|  |  |  |  |  |  |  |  |  |  | lumefantrine<br>concentration |  |  |  |
| 18 | DP | EFV | NVP | 159<sup>1</sup> | 8 | 61 | 0 | 42 | RD<sup>2</sup> | Univariable | 5.0% | 1.6-8.4 | 0.074 |

<sup>1</sup> excludes 1 early treatment failure
<sup>2</sup> risk difference (RD) calculated from proportions, not provided in the publication. Odds ratio (OR) cannot be calculated due to 0 events in one group.
ACT: artemisinin combination therapy; AL: artemether-lumefantrine; ART: antiretroviral therapy; DP: dihydroartemisinin-piperaquine; EFV: efavirenz; HR: hazard ratio; LPV/r: lopinavir/ritonavir; N/A: information not available; NVP: nevirapine; OR: odds ratio; RR: relative risk

### Early parasitological response

Four studies (ID 7, ID 11, ID 12, ID 19) compared parasite clearance between PLHIV and HIV-uninfected patients (**Table 5**). In a study where patients were treated with AL or SP (ID 12), no significant difference in prevalence of parasite positive readings were found on day 3 (24), while in two studies (ID 7, ID 11) with patients treated with AL or DP, a significantly faster clearance was observed in HIV-uninfected patients (34, 35). In study ID 11, median parasite clearance half-life was larger by 25% in PLHIV compared to HIV-uninfected patients (3.5 hours versus 2.8 hours) and this was consistent across the three HIV treatment groups (EFV, NVP and LPV/r) (34). In study ID 19, Chijioke-Nwauche *et al*. showed similar proportion of patients with parasite positivity on day 3 between groups, but baseline parasitaemia, even though detected by microscopy, was undetectable by nested-PCR in 78.1% of patients (27).

**Table 5.**
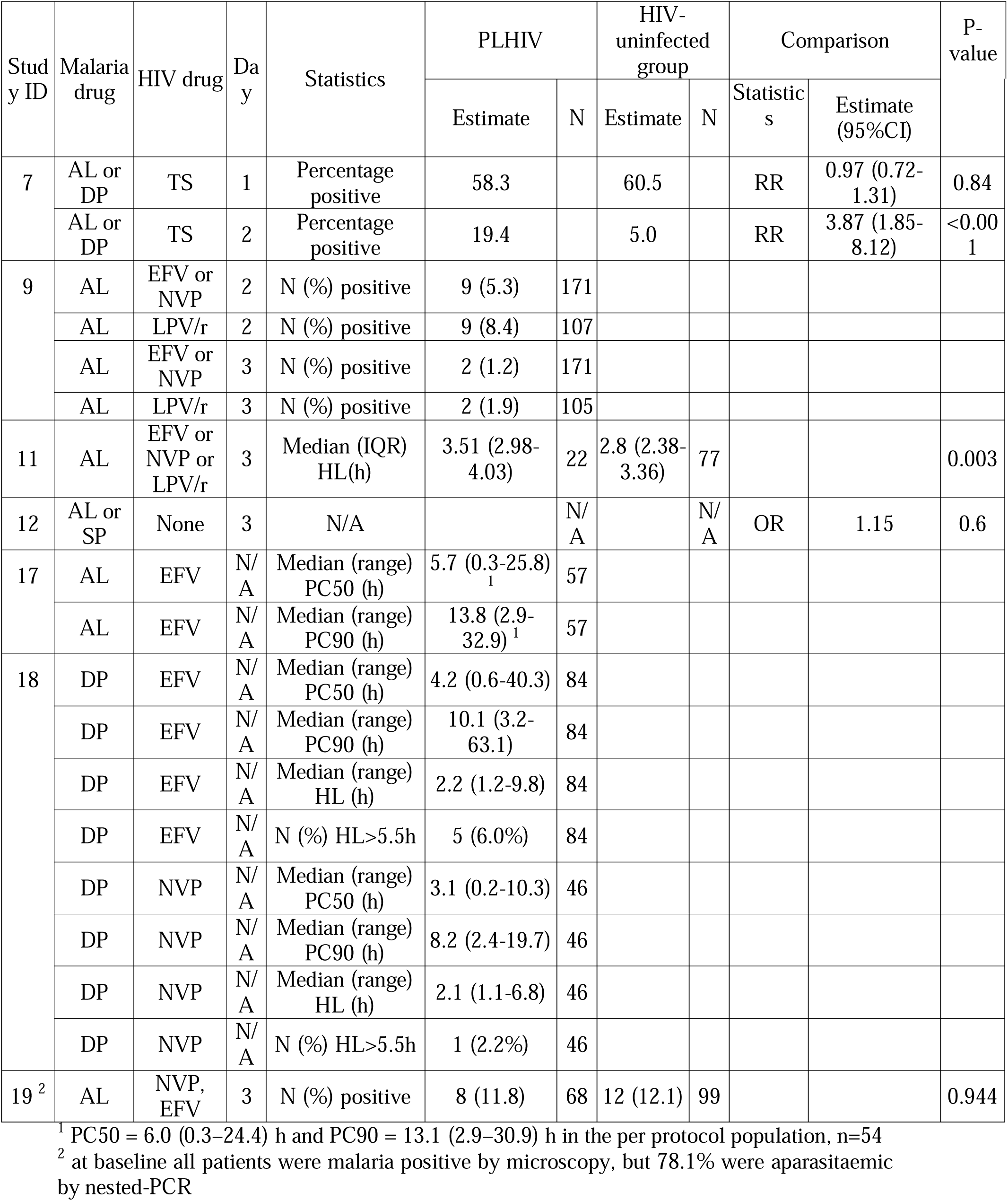
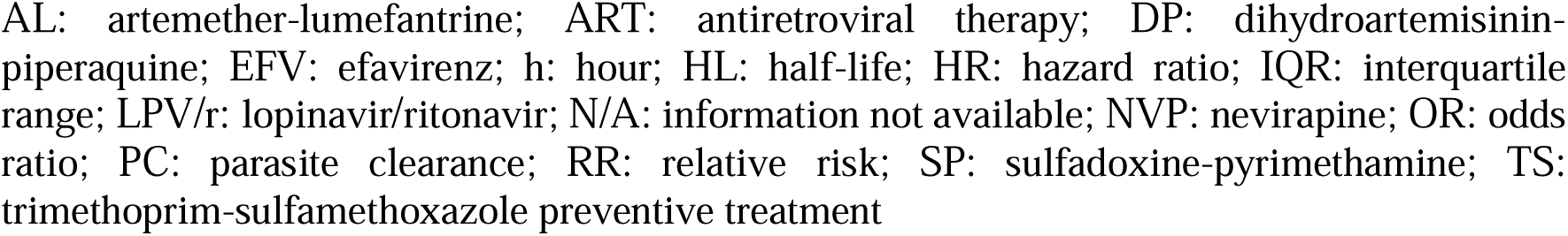
Summary of reported results for early parasitological response.

| Study ID | Malaria drug | HIV drug | Day | Statistics | PLHIV |  | HIV-uninfected group |  | Comparison |  | P-value |
| --- | --- | --- | --- | --- | --- | --- | --- | --- | --- | --- | --- |
|  |  |  |  |  | Estimate | N | Estimate | N | Statistics | Estimate (95%CI) |  |
| 7 | AL or DP | TS | 1 | Percentage positive | 58.3 |  | 60.5 |  | RR | 0.97 (0.72-1.31) | 0.84 |
|  | AL or DP | TS | 2 | Percentage positive | 19.4 |  | 5.0 |  | RR | 3.87 (1.85-8.12) | <0.001 |
| 9 | AL | EFV or NVP | 2 | N (%) positive | 9 (5.3) | 171 |  |  |  |  |  |
|  | AL | LPV/r | 2 | N (%) positive | 9 (8.4) | 107 |  |  |  |  |  |
|  | AL | EFV or NVP | 3 | N (%) positive | 2 (1.2) | 171 |  |  |  |  |  |
|  | AL | LPV/r | 3 | N (%) positive | 2 (1.9) | 105 |  |  |  |  |  |
| 11 | AL | EFV or NVP or LPV/r | 3 | Median (IQR) HL(h) | 3.51 (2.98-4.03) | 22 | 2.8 (2.38-3.36) | 77 |  |  | 0.003 |
| 12 | AL or SP | None | 3 | N/A |  | N/A |  | N/A | OR | 1.15 | 0.6 |
| 17 | AL | EFV | N/A | Median (range) PC50 (h) | 5.7 (0.3-25.8)<sup>1</sup> | 57 |  |  |  |  |  |
|  | AL | EFV | N/A | Median (range) PC90 (h) | 13.8 (2.9-32.9)<sup>1</sup> | 57 |  |  |  |  |  |
| 18 | DP | EFV | N/A | Median (range) PC50 (h) | 4.2 (0.6-40.3) | 84 |  |  |  |  |  |
|  | DP | EFV | N/A | Median (range) PC90 (h) | 10.1 (3.2-63.1) | 84 |  |  |  |  |  |
|  | DP | EFV | N/A | Median (range) HL (h) | 2.2 (1.2-9.8) | 84 |  |  |  |  |  |
|  | DP | EFV | N/A | N (%) HL>5.5h | 5 (6.0%) | 84 |  |  |  |  |  |
|  | DP | NVP | N/A | Median (range) PC50 (h) | 3.1 (0.2-10.3) | 46 |  |  |  |  |  |
|  | DP | NVP | N/A | Median (range) PC90 (h) | 8.2 (2.4-19.7) | 46 |  |  |  |  |  |
|  | DP | NVP | N/A | Median (range) HL (h) | 2.1 (1.1-6.8) | 46 |  |  |  |  |  |
|  | DP | NVP | N/A | N (%) HL>5.5h | 1 (2.2%) | 46 |  |  |  |  |  |
| 19<sup>2</sup> | AL | NVP, EFV | 3 | N (%) positive | 8 (11.8) | 68 | 12 (12.1) | 99 |  |  | 0.944 |

<sup>1</sup> PC50 = 6.0 (0.3–24.4) h and PC90 = 13.1 (2.9–30.9) h in the per protocol population, n=54
<sup>2</sup> at baseline all patients were malaria positive by microscopy, but 78.1% were aparasitaemic by nested-PCR
AL: artemether-lumefantrine; ART: antiretroviral therapy; DP: dihydroartemisinin-piperaquine; EFV: efavirenz; h: hour; HL: half-life; HR: hazard ratio; IQR: interquartile range; LPV/r: lopinavir/ritonavir; N/A: information not available; NVP: nevirapine; OR: odds ratio; PC: parasite clearance; RR: relative risk; SP: sulfadoxine-pyrimethamine; TS: trimethoprim-sulfamethoxazole preventive treatment

Three further studies (ID 9, ID 17, ID 18) reported the results for early parasitological response in PLHIV only, either on EFV or NVP treatment. Parasite clearance was faster in PLHIV receiving EFV and treated with AL, compared to those treated with DP; and within the DP treatment group (ID 18), PLHIV receiving NVP parasite clearance was faster compared to PLHIV receiving EFV, although formal comparison was not presented (32). Parasite positivity on day 2 or 3 was similar between patients on NVP or EFV, compared to LPV/r in study ID 9 (33).

### Gametocytaemia

No quantitative analysis was possible. Three studies (ID 6, ID 11, ID 12) compared gametocyte carriage between PLHIV co-infected with malaria and HIV-uninfected patients. In study ID 6, a higher risk of developing gametocytes within 28 days follow-up was found in patients with TS prophylaxis versus no TS, RR=1.76 (95%CI 1.29-2.40, p<0.001) (36). In the same study, after adjusting for TS, patients’ age, treatment arm and recurrent parasitaemia status, the risk was not different between PLHIV and HIV-uninfected patients, RR=1.29 (95% CI 0.74-2.24, p=0.373) (36). TS prophylaxis was also associated with delayed gametocyte clearance, with HR=1.32 (95%CI 1.05–1.64, p=0.02) compared to patients without TS prophylaxis.

In study ID 11, the proportion of episodes in which patients developed gametocytes after treatment (days 1-28) was 17% (of 188 episodes) in HIV-uninfected group compared to 20% (of 70 malaria episodes), 29.6% (62 malaria episodes) and 36% (in 50 episodes) in LPV/r, NVP, EFV treated groups of PLHIV (p=0.008 for comparison between HIV-uninfected and a combined group treated with NVP or EFV) (26) respectively. In the study ID 12, presence of gametocytes on days 3, 7, 14, 28, 45 was not significantly different between HIV-uninfected patients and PLHIV not yet on treatment (24).

The two ART regimens (NVP or EFV, LPV/r) in the study ID 9 were not different in respect to gametocyte carriage (defined as the appearance of gametocytes on days 2-28 among those without gametocytes on day 0): 8.3% (n=12/145) compared to 6.1% (n=6/99), respectively, although the pattern were consistent with results of another study of lower gametocytes carriage in LPV/r group (33).

### Pharmacokinetic properties

#### Lumefantrine and desbutyl-lumefantrine

Eleven studies provided lumefantrine concentrations measured on day 7 for different ART regimens (**Additional File 7, Fig. 2**). Highest day 7 concentrations were observed in patients treated with LPV/r (**Fig. 2**). The pooled estimate of the weighted ratio between lumefantrine concentration geometric mean in patients treated with LPV/r compared to EFV was 7.89 (95%CI 6.57-9.50, p<0.001, I^2^=77.6%, chi-square test for heterogeneity p=0.011, 3 studies); and between LPV/r and NVP it was 2.83 (95%CI 2.34-3.41, p<0.001, I^2^=34.8%, chi-square test for heterogeneity p=0.216, 3 studies).

**Fig. 2.**
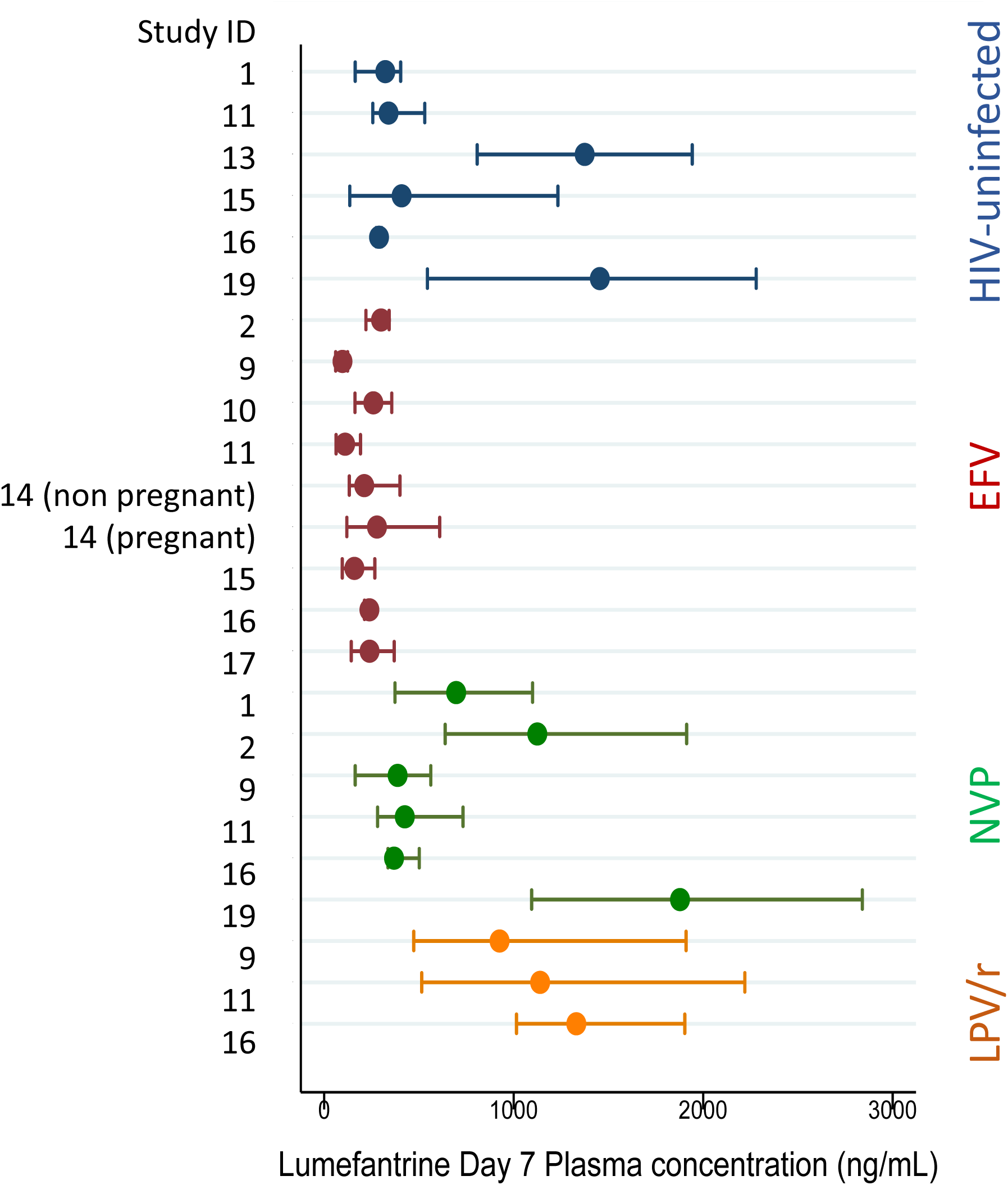
Day 7 lumefantrine concentration in PLHIV treated with different antiretroviral therapies and in HIV-uninfected patients. EFV: efavirenz-based antiretroviral therapy; NVP: nevirapine-based antiretroviral therapy; LPV/r: lopinavir/ritonavir-based antiretroviral therapy. Study reported central tendency measure (mean or median or geometric mean) and estimated interquartile range are presented (for details of calculation see methods).

Lowest day 7 lumefantrine concentrations were observed in patients treated with EFV. The pooled weighted geometric mean ratio of lumefantrine concentration comparing NVP and EFV was estimated as 2.37 (95%CI 2.05-2.73, p<0.001, I^2^=95.1%, chi-square test for heterogeneity p<0.001, 3 studies).

Day 7 lumefantrine concentration in patients treated with NVP was also higher compared to HIV-uninfected patients with a pooled ratio of geometric mean 1.31 (95%CI 1.16-1.47, p<0.001, I^2^=27.6%, chi-square test for heterogeneity p=0.246, 4 studies), while patients treated with EFV had lower concentrations compared to HIV-uninfected patients with a pooled ratio of geometric mean 0.76 (95%CI 0.81-0.70, p<0.001, I^2^=96.2%, chi-square test for heterogeneity p<0.001, 3 studies).

Studies (**Table 6**) which reported lumefantrine elimination half-life in PLHIV showed that lumefantrine clearance was faster when treated with EFV compared to other ART regimens (ID 11), to TS prophylaxis only (ID 10) or not yet on treatment (ID 15). Lumefantrine half-life was shorter by 30-60% (median [IQR] 23.7h [21.8–46.0] vs. 64.3h [52.0–120.6], p<0.0001 in study ID 11 (26): the geometric mean was [90%CI] 59.2h [46.7, 75.1] vs. 89.5h [75.3, 106.3], p=0.033 in study ID 15 (28), and the mean was [95% CI] 33h [30.9-35.7] vs. 36 h [34.1-37.9], p=0.036 in study ID 10 (37).

**Table 6.** Summary of the reported lumefantrine AUC and elimination half-life.

| Study ID | Statistics | HIV-uninfected group | TS prophylaxis | NVP | EFV | LPV/r | ATV/r | PLHIV not yet on ART |
| --- | --- | --- | --- | --- | --- | --- | --- | --- |
| **Lumefantrine half-life (h)** |  |  |  |  |  |  |  |  |
| 10 | Mean (95%CI) |  | 36.0 (34.1-37.9) | 33.3 (30.9-35.7) | 33.3 (30.9-35.7) |  |  |  |
| 11 | Median (IQR) |  |  | 63.4 (46.8-111.1) | 23.7 (21.8-46.0) | 98.7 (88.4-119.1) |  | 64.3 (52.0-120.6) |
| 13 | Mean (SEM) | 31.16 (1.86) |  |  |  |  | 42.59 (3.77) |  |
| 15 | GM (90%CI) |  |  |  | 59.2 (46.7-75.1) |  |  | 89.5 (75.3-106.3) |
| **Lumefantrine AUC (ng.h/ml)** |  |  |  |  |  |  |  |  |
| 2<sup>1</sup> | Median (IQR), AUC<sub>0-∞</sub> |  |  | 977645 (688477-1383975) | 303130 (211080-431962) |  |  | 784830 (547405-1116250) |
| 10 | Mean (95%CI) AUC<sub>0-∞</sub> |  | 375200 (349700-400700) | 264800 (243100-286500) | 264800 (243100-286500) |  |  |  |
| 11 | GM (90%CI), AUC<sub>0-∞</sub> |  |  | 278000 (228000-339000) | 130000 (107000-157000) | 579000 (477000 - 704000) |  | 270000 (232000-313000) |
| 13 | Mean (SEM), AUC<sub>0-168</sub> | 447976 (±80887) |  |  |  |  | 670530 (±157173) |  |
| 15 | GM (90%CI), AUC<sub>0-∞</sub> |  |  |  | 188 (125-281) |  |  | 287 (237-349) |
| 16 | Mean (SEM), AUC<sub>0-168</sub> | 83508 (±5361) |  | 125285 (±35221) | 58396 (±8019) | 357295 (±5156) |  |  |

<sup>1</sup> Population PK study, values presented are from 9960 simulations
ART: antiretroviral therapies; ATV/r: atazanavir-ritonavir; AUC: area under the curve (from 0 hour to infinite or to 168 hours); CI: confidence interval (90% or 95%); EFV: efavirenz; IQR: interquartile range; GM: geometric mean; LPV/r: lopinavir/ritonavir; NVP: nevirapine; PLHIV: people living with HIV; SEM: standard error of the mean; TS: trimethoprim-sulfamethoxazole preventive treatment

Area under the curve (AUC) comparison between patients on different ART regimens or TS prophylaxis was reported in 6 studies (**Table 6**). Compared to HIV-uninfected patients (ID 16) or to PLHIV not yet on treatment (ID 11), increased AUC was observed in PLHIV treated with LPV/r (ratio 4.3 and 2.1, respectively). AUC was decreased in PLHIV treated with EFV compared to those not yet on treatment (ratio 0.5 in study ID 11 and 0.7 in study ID 15), and similar in patients treated with NVP (ratio 1, ID 11). Also, simulations after the population pharmacokinetic modelling in study ID 2 estimated the ratio of median lumefantrine AUC for EFV-treated and NVP-treated patients to the PLHIV but ART-naïve to be 0.4 and 1.2, respectively (38).

#### Piperaquine

Only one study (ID 4) assessed the pharmacokinetics of piperaquine in PLHIV with uncomplicated *P. falciparum* malaria. In the population pharmacokinetic model of 218 malaria episodes in children from Uganda, no evidence of a significant effect on any clearance or volume distribution parameters was observed for prophylactic treatment with TS (n=41), PLHIV not yet on ART (n=12) nor antiretroviral therapy (n=10) (39). No quantitative results were provided.

#### Artemether and dihydroartemisinin (DHA)

Three studies (ID 1, ID 11, ID 15) that investigated pharmacokinetic parameters in PLHIV also included HIV-uninfected patients; these studies evaluated artemether and DHA AUC, maximum concentration (C_max_) and time to maximum concentration (T_max_). For both artemether and DHA, C_max_ and AUC_0-8_ were consistently lower in PLHIV compared to uninfected patients (**Table 7**), although the differences were not significant for the LPV/r group. The ratio of C_max_ and AUC _0-8_ after the first and last dose was studied in a subset of patients in the study by Kajubi *et al.* (34), and while the decrease in geometric means for artemether was observed for HIV-uninfected patients as well as for LPV/r and NVP but not EFV-treated PLHIV, the increase in values for DHA were only observed in HIV-uninfected patients and not in patients on ART.

**Table 7.** Effect of various ART regimens on DHA and artemether concentration levels, measured after the last AL dose.

|  | Ratio (95% CI) of the geometric mean for PLHIV on various ART regimens to the geometric mean for HIV-uninfected patients |  |  |
| --- | --- | --- | --- |
|  | EFV* | NVP** | LPV/r*** |
| Artemether AUC<sub>0-8</sub> | 0.43 (0.30 - 0.62)<sup>a</sup> | 0.34 (0.25-0.46)<sup>1</sup> | 0.75 (0.53-1.05) |
| Artemether C<sub>max</sub> | 0.46 (0.30 - 0.69)<sup>b</sup> | 0.34 (0.23-0.45)<sup>2</sup> | 0.75 (0.50-1.11) |
| DHA AUC<sub>0-8</sub> | 0.37 (0.28-0.51)<sup>c</sup> | 0.70 (0.55-0.88)<sup>3</sup> | 0.81 (0.59-1.10) |
| DHA C<sub>max</sub> | 0.40 (0.28 - 0.58)<sup>d</sup> | 0.66 (0.50-0.87)<sup>4</sup> | 0.83 (0.57-1.19) |

\* shows results for the combined studies (ID 11, ID 15)
\*\* shows results for the combined studies (ID 1, ID 11)
\*\*\* shows results from study ID 11
<sup>a</sup> I-squared = 0.0%, p-value for heterogeneity = 0.150
<sup>b</sup> I-squared=0.0%, p-value for heterogeneity = 0.474
<sup>c</sup> I-squared=81.0%, p-value of heterogeneity = 0.022
<sup>d</sup> I-squared = 0.0%, p-value for heterogeneity = 0.322
<sup>1</sup> I-squared = 51.7%, p-value for heterogeneity = 0.150
<sup>2</sup> I-squared=43.0%, p-value for heterogeneity = 0.185
<sup>3</sup> I-squared=0.0%, p-value of heterogeneity = 0.445
<sup>4</sup> I-squared = 0.0%, p-value for heterogeneity = 0.842
ART: antiretroviral therapies; AUC: area under the curve (from 0 hour to 8 hours); CI: confidence interval (95%); C<sub>max</sub>: maximum concentration; DHA: dihydroartemisinin; EFV: efavirenz; LPV/r: lopinavir/ritonavir; CI: confidence interval; PLHIV: people living with HIV

No differences in time to maximum DHA concentration were detected between PLHIV treated with NVP, EFV or LPV/r compared to HIV-uninfected patients (26, 40). For artemether, in one study (ID 1), T_max_ was observed significantly earlier (p=0.028) in NVP (1.3 h) compared to historical controls (2h) which was not in agreement with findings of the other study (ID 11), where a trend of rather later T_max_ (median 2.1 NVP, EVP and 3.0 in LPV/r) in PLHIV was observed compared to HIV-uninfected (2h).

#### Assessment of bias

The risk of bias in individual studies was generally considered to be low to moderate across all the domains considered for seven RCTs (**Additional Files 3 and 4**). In all RCTs, assessment of outcome (parasite or gametocyte positivity, drug concentration) was blinded as it was evaluated in the laboratory, independently from the clinical team. Of the 12 non-randomized studies, 11 were considered to be at low-moderate risk of bias across participant selection, intervention classification, selective reporting and outcome measurement domains. Only 3 studies were considered to be at low risk of bias due to confounding, while the remaining 9 studies were at moderate-high risk.

#### Certainty of evidence

There were a couple of limitations in the few studies conducted. Firstly, a comparison of the risk of recurrence between PLHIV on TS prophylaxis and HIV-uninfected patients was performed based on total number of patients in each treatment arm and ignoring the losses to follow-up. Secondly, comparison of lumefantrine day 7 concentrations was based on the mean and standard error of the log-transformed concentration values, which for many studies were estimated from median and interquartile range. However, for three analyses for which evidence synthesis was possible, indirectness, imprecision and inconsistency were classified as not serious. The publication bias is low possibly because of the limited number of studies being conducted in patients co-infected with malaria and HIV. When all these assessments are taken together, the certainty of evidence generated from this review is likely to be low-moderate based on the GRADE guidelines (18).

## Discussion

The main finding of this systematic review is the paucity and heterogeneity of studies set in Africa comparing the efficacy of ACT in PLHIV, either untreated, on TS prophylaxis or under different ART regimens, with HIV-uninfected persons.

The only study (ID 12) looking at malaria efficacy after receiving AL or SP in PLHIV but treatment-naive adult patients (24) didn’t allow for the evaluation of efficacy of individual anti-malarial drug and failed to show any differences between PLHIV and HIV-uninfected groups in terms of parasite and gametocyte clearance, malaria recrudescence, recurrence, or reinfection on day 45. A UNAIDS report from 2022 estimated that 78% of PLHIV receive HIV treatment in sub-Saharan Africa with substantial differences in access to treatment within countries, including in children living with HIV (41). It is however unlikely to see new studies looking at anti-malarial drug efficacy in PLHIV ART-naive, as the latest WHO guidelines encourage rapid treatment of newly diagnosed HIV patients (42). Therefore, delaying ART initiation until completion of malaria treatment, usually day 28 or day 42 (14) may be unethical.

Seven of the eleven studies reporting late treatment failure included HIV-uninfected participants. Risk of malaria recurrence was the most often reported outcome and the results were different for children and adults. In Uganda, where TS prophylaxis has been shown to reduce the risk of new malaria infections (43, 44), risk of recurrence was lower in children living with HIV on TS prophylaxis (with or without ART regimen) when receiving AL or ASAQ for their malaria episode (29, 30, 45). Further studies conducted in Uganda have found that HIV-uninfected children receiving AL had significantly increased odds of malaria recurrence on day 28 compared to children living with HIV under any of the 3 ART regimens proposed (LPV/r, NVP, or EFV), (26). The two studies conducted in adults (one each in Uganda and Nigeria) reported higher proportion (statistically non-significant) of recurrence in PLHIV on EFV or NVP compared to HIV-uninfected patients. These findings are in line with previous reviews reporting that differences in response to treatment against malaria in adults living with HIV in endemic areas may be explained by impaired acquired immunity (8, 46).

A longer parasite clearance was observed in Ugandan children when AL or DP was administered in the presence of EFV, NVP, LPV/r or TS (34, 35), a finding consistent with another report showing that children living with HIV have slower parasite clearance compared to those HIV-uninfected (34, 35, 47). Additionally, the proportion of children living with HIV treated with LPV/r, EFV or NVP who developed gametocytes was higher than that in HIV-uninfected patients. An increased gametocyte carriage coupled with a slower parasite clearance in PLHIV on ART regimen is concerning as this population may serve as an unwitting reservoir for transmission (48) and subsequent spread of resistant parasites (49). Better understanding the role of a weakened immune status due to HIV infection in the emergence of anti-malarial drug-resistant parasites is one of the recommendations suggested by the WHO in its strategic report to tackle this threat (50).

Drug-drug interaction between LPV/r-, NVP-, or EFV-based regimen and AL has been described in PLHIV without malaria (10, 51, 52) or in healthy adults (53). Both nevirapine and efavirenz induce the cytochrome P450 enzyme system, which metabolizes artemether and lumefantrine, while lopinavir and ritonavir inhibit the system, which may result in an increased artemether and lumefantrine plasma concentration. Pharmacokinetic data extracted from studies considered in this review confirms these results. Comparison of different ART regimens showed a 3-fold increased risk of recurrence among children living with HIV on NVP or EFV compared to those on LPV/r, but also showed this effect disappeared once day 7 lumefantrine concentration was accounted in the analysis. Day 7 lumefantrine concentrations were highest in patients treated with LPV/r and lowest in patients treated with EFV, including when compared to HIV-uninfected patients (two studies) or PLHIV not yet on ART (one study). In addition, for PLHIV on EFV-based regimen, clearance of lumefantrine was significantly faster compared to that in HIV-uninfected patients; therapeutic lumefantrine concentration level of 280 ng/mL was not reached by half of the patients in one study (ID 14) and the median or mean plasma concentration level in 3 out of the 4 other studies was below that threshold. PLHIV on NVP had a similar AUC and a higher lumefantrine plasma concentration on day 7 compared to that of HIV-uninfected patients and a median concentration above the therapeutic level. PLHIV treated with a standard 3-day anti-malarial treatment for uncomplicated *P. falciparum* malaria may contribute between 2.6-6.6% of estimated excess failures because of suboptimal anti-malarial drug dosing (3). Lower exposure to lumefantrine, especially in PLHIV on EFV is of concern as it could not only result in treatment failure but also contribute to the spread of resistant parasites. The poor efficacy of AL in PLHIV on EFV has already been described (54, 55). Prolonging the duration of the AL treatment from 3 to 5 days could be considered as it would reduce the risk of having a day 7 lumefantrine plasma concentration below the therapeutic threshold (55). Malaria patients co-infected with HIV are considered a special risk group in the WHO malaria treatment guidelines (12); however, to date, no dose adjustment for this risk group has been officially endorsed. The WHO now recommends the use of dolutegravir-based ART regimen as first line treatment for PLHIV and EFV-based regimen as an alternative first-line regimen; dolutegravir has fewer drug-drug interactions and does not seem to modify AL pharmacokinetics (56, 57).

This review is limited by the minimal meta-analyses performed due to differences in the presentation of the data and the reported estimates.

## Conclusion

Limited data on ACT treatment outcomes or drug exposure in PLHIV in Africa remains a reality to date, and there is important heterogeneity in study designs limiting the interpretation of the results. PLHIV on EFV appears nevertheless to be at risk of suboptimal dosing when treated with a standard 3-day AL regimen for uncomplicated *P. falciparum* malaria and also at a higher risk of treatment failure. Conducting an individual patient data meta-analysis to explore the impact of antiretroviral therapy on anti-malarial treatment would help understand these complex interactions better.

## Supporting information

Suppl_File_preprint

## Data Availability

All data generated or analysed during this study are included in this manuscript and its supplementary information files

## List of abbreviations

ACT: artemisinin-based combination therapy
AL: artemether-lumefantrine
AP: artesunate-pyronaridine
ART: antiretroviral therapy
ASAQ: artesunate-amodiaquine
ASMQ: artesunate-mefloquine
ASSP: artesunate-sulfadoxine-pyrimethamine
ATVr: atazanavir-ritonavir
AUC: area under the curve
CI: confidence interval
C_max_: maximum concentration
DHA: dihydroartemisinin
DP: dihydroartemisinin-piperaquine
EFV: efavirenz
HAART: highly active antiretroviral therapy
HIV: human immunodeficiency virus
HR: Hazard Ratio
IQR: interquartile range
KM: Kaplan-Meier
LPV/r: lopinavir-ritonavir
LUM: lumefantrine
N/A: non available
NVP: nevirapine
OR: Odds Ratio
PCR: polymerase chain reaction
PK: pharmacokinetics
PLHIV: People living with HIV
PPQ: piperaquine
RCT: randomized clinical trial
RR: risk ratio
SP: sulfadoxine-pyrimethamine
T_max_: time to maximum concentration
TS: trimethoprim-sulfamethoxazole
WHO: World Health Organization
WWARN: WorldWide antimalarial resistance network

## Declarations

### Ethics approval and consent to participate

Not applicable.

### Consent for publication

Not applicable.

### Availability of data and materials

All data generated or analysed during this study are included in this published article and its supplementary information files.

### Authors’ contributions

Conceptualization: AT, PG, KS. Investigation: AT, AS, MP, EH. Data curation: AT, AS, MP, KS. Formal analysis: AT, AS, PD, KS. Supervision: KB, PD, PG, AT, AS, MP, KS. Writing original draft: AT, AS, PD, KB, KS, VIC. All authors read and approved the final manuscript.

## Acknowledgements

Not applicable.

## Authors’ information

Kasia Stepniewska□ and Verena Carrara have contributed equally to the work.

## Additional Files

Additional File 1. Search terms for the original search in MEDLINE, EMBASE, Web of Science (all Databases), Cochrane Central, WHO Global Index Medicus and Clinicaltrials.gov (Word.docx)

Additional File 2. Search terms for the subsequent searches in the WWARN Clinical Trial Library (Word.docx)

Additional File 3. ROB assessment for non-RCTs: methods and outline of the risk of bias tables and responses to each of the signalling questions (Excel.xlsx)

Additional File 4. ROB assessment for RCTs: methods and outline of the risk of bias tables and responses to each of the signalling questions (Excel.xlsx)

Additional File 5. Inclusion criteria for enrolment of people living with HIV (PLHIV) in each study included in the review and their baseline CD4 measurements (Word.docx)

Additional File 6. Geographical distribution of the 19 study sites and their malaria transmission level as reported in individual manuscripts (Figure.tiff)

Additional File 7. Summary of the reported day 7 lumefantrine concentrations (ng/mL) (Word.docx)

## References

1. AIDSinfo. Global data on HIV epidemiology and response 2023 [cited 2025 January, 27]. Available from: http://aidsinfo.unaids.org/.

2. WHO. World malaria report 2024: addressing inequity in the global malaria response. Geneva: World Health Organization; 2024.

3. Takyi A, Carrara VI, Dahal P, Przybylska M, Harriss E, Insaidoo G, et al. Characterisation of populations at risk of sub-optimal dosing of artemisinin-based combination therapy in Africa. PLoS Glob Public Health. 2023;3:e0002059.

4. Herrero MD, Rivas P, Rallon NI, Ramirez-Olivencia G, Puente S. HIV and malaria. AIDS Rev. 2007;9:88–98.

5. Kamya MR, Gasasira AF, Yeka A, Bakyaita N, Nsobya SL, Francis D, et al. Effect of HIV-1 infection on antimalarial treatment outcomes in Uganda: a population-based study. J Infect Dis. 2006;193:9–15.

6. Cohen C, Karstaedt A, Frean J, Thomas J, Govender N, Prentice E, et al. Increased prevalence of severe malaria in HIV-infected adults in South Africa. Clin Infect Dis. 2005;41:1631–7.

7. ter Kuile FO, Parise ME, Verhoeff FH, Udhayakumar V, Newman RD, van Eijk AM, et al. The burden of co-infection with human immunodeficiency virus type 1 and malaria in pregnant women in sub-saharan Africa. Am J Trop Med Hyg. 2004;71(2 Suppl):41–54.

8. Flateau C, Le Loup G, Pialoux G. Consequences of HIV infection on malaria and therapeutic implications: a systematic review. Lancet Infect Dis. 2011;11:541–56.

9. Autran B, Carcelain G, Li TS, Blanc C, Mathez D, Tubiana R, et al. Positive effects of combined antiretroviral therapy on CD4+ T cell homeostasis and function in advanced HIV disease. Science. 1997;277:112–6.

10. Kredo T, Mauff K, Workman L, Van der Walt JS, Wiesner L, Smith PJ, et al. The interaction between artemether-lumefantrine and lopinavir/ritonavir-based antiretroviral therapy in HIV-1 infected patients. BMC Infect Dis. 2016;16:30.

11. Nsanzabana C, Rosenthal PJ. In vitro activity of antiretroviral drugs against Plasmodium falciparum. Antimicrob Agents Chemother. 2011;55:5073–7.

12. WHO. Guidelines for malaria, 3 June 2022. Geneva: World Health Organization; 2022.

13. Takata J, Sondo P, Humphreys GS, Burrow R, Maguire B, Hossain MS, et al. The WorldWide Antimalarial Resistance Network Clinical Trials Publication Library: a live, open-access database of Plasmodium treatment efficacy trials. Am J Trop Med Hyg. 2020;103:359–68.

14. WHO. Methods for surveillance of antimalarial drug efficacy. Geneva: World Health Organization; 2009.

15. WHO. Methods and techniques for clinical trials on antimalarial drug efficacy: genotyping to identify parasite populations. Geneva: World Health Organization; 2008.

16. Sterne JAC, Savovic J, Page MJ, Elbers RG, Blencowe NS, Boutron I, et al. RoB 2: a revised tool for assessing risk of bias in randomised trials. BMJ. 2019;366:l4898.

17. Sterne JA, Hernan MA, Reeves BC, Savovic J, Berkman ND, Viswanathan M, et al. ROBINS-I: a tool for assessing risk of bias in non-randomised studies of interventions. BMJ. 2016;355:i4919.

18. Guyatt GH, Oxman AD, Sultan S, Glasziou P, Akl EA, Alonso-Coello P, et al. GRADE guidelines: 9. Rating up the quality of evidence. J Clin Epidemiol. 2011;64:1311–6.

19. Klein JP, Logan B, Harhoff M, Andersen PK. Analyzing survival curves at a fixed point in time. Stat Med. 2007;26:4505–19.

20. Hozo SP, Djulbegovic B, Hozo I. Estimating the mean and variance from the median, range, and the size of a sample. BMC Med Res Methodol. 2005;5:13.

21. VassarStats website for statistical computation. Estimation of a sample’s mean and variance from its median and range [cited 2023 December, 27]. Available from: http://vassarstats.net/median_range.html.

22. Wan X, Wang W, Liu J, Tong T. Estimating the sample mean and standard deviation from the sample size, median, range and/or interquartile range. BMC Med Res Methodol. 2014;14:135.

23. Battistini Garcia SA, Zubair M, Guzman N. CD4 cell count and HIV. [Updated 2025 Jan 19]. In: StatPearls [Internet]. Treasure Island (FL): StatPearls Publishing; 2025. Available from: https://www.ncbi.nlm.nih.gov/books/NBK513289/.

24. Van Geertruyden JP, Mulenga M, Mwananyanda L, Chalwe V, Moerman F, Chilengi R, et al. HIV-1 immune suppression and antimalarial treatment outcome in Zambian adults with uncomplicated malaria. J Infect Dis. 2006;194:917–25.

25. Maganda BA, Minzi OM, Kamuhabwa AA, Ngasala B, Sasi PG. Outcome of artemether-lumefantrine treatment for uncomplicated malaria in HIV-infected adult patients on anti-retroviral therapy. Malar J. 2014;13:205.

26. Parikh S, Kajubi R, Huang L, Ssebuliba J, Kiconco S, Gao Q, et al. Antiretroviral choice for HIV impacts antimalarial exposure and treatment outcomes in Ugandan children. Clin Infect Dis. 2016;63:414–22.

27. Chijioke-Nwauche I, van Wyk A, Nwauche C, Beshir KB, Kaur H, Sutherland CJ. HIV-positive nigerian adults harbor significantly higher serum lumefantrine levels than HIV-negative individuals seven days after treatment for Plasmodium falciparum infection. Antimicrob Agents Chemother. 2013;57:4146–50.

28. Hughes E, Mwebaza N, Huang L, Kajubi R, Nguyen V, Nyunt MM, et al. Efavirenz-based antiretroviral therapy reduces artemether-lumefantrine exposure for malaria treatment in HIV-infected pregnant women. J Acquir Immune Defic Syndr. 2020;83:140–7.

29. Verret WJ, Arinaitwe E, Wanzira H, Bigira V, Kakuru A, Kamya M, et al. Effect of nutritional status on response to treatment with artemisinin-based combination therapy in young Ugandan children with malaria. Antimicrob Agents Chemother. 2011;55:2629–35.

30. Wanzira H, Kakuru A, Arinaitwe E, Bigira V, Muhindo MK, Conrad M, et al. Longitudinal outcomes in a cohort of Ugandan children randomized to artemether-lumefantrine versus dihydroartemisinin-piperaquine for the treatment of malaria. Clin Infect Dis. 2014;59:509–16.

31. Kakuru A, Achan J, Muhindo MK, Ikilezi G, Arinaitwe E, Mwangwa F, et al. Artemisinin-based combination therapies are efficacious and safe for treatment of uncomplicated malaria in HIV-infected Ugandan children. Clin Infect Dis. 2014;59:446–53.

32. Sevene E, Banda CG, Mukaka M, Maculuve S, Macuacua S, Vala A, et al. Efficacy and safety of dihydroartemisinin-piperaquine for treatment of Plasmodium falciparum uncomplicated malaria in adult patients on antiretroviral therapy in Malawi and Mozambique: an open label non-randomized interventional trial. Malar J. 2019;18:277.

33. Achan J, Kakuru A, Ikilezi G, Ruel T, Clark TD, Nsanzabana C, et al. Antiretroviral agents and prevention of malaria in HIV-infected Ugandan children. N Engl J Med. 2012;367:2110–8.

34. Kajubi R, Huang L, Were M, Kiconco S, Li F, Marzan F, et al. Parasite clearance and artemether pharmacokinetics parameters over the course of artemether-lumefantrine treatment for malaria in human immunodeficiency virus (HIV)-infected and HIV-uninfected Ugandan children. Open Forum Infect Dis. 2016;3:ofw217.

35. Muhindo MK, Kakuru A, Jagannathan P, Talisuna A, Osilo E, Orukan F, et al. Early parasite clearance following artemisinin-based combination therapy among Ugandan children with uncomplicated Plasmodium falciparum malaria. Malar J. 2014;13:32.

36. Kakuru A, Jagannathan P, Arinaitwe E, Wanzira H, Muhindo M, Bigira V, et al. The effects of ACT treatment and TS prophylaxis on Plasmodium falciparum gametocytemia in a cohort of young Ugandan children. Am J Trop Med Hyg. 2013;88:736–43.

37. Musoke D, Bergmann TK, Ntale M, Sodemann M, Ogwal-Okeng J. Pharmacokinetics of lumefantrine in adults co-infected with malaria and HIV-1: with and without efavirenz-based antiretroviral therapy. Int J Trop Med. 2012;7:187–92.

38. Maganda BA, Ngaimisi E, Kamuhabwa AA, Aklillu E, Minzi OM. The influence of nevirapine and efavirenz-based anti-retroviral therapy on the pharmacokinetics of lumefantrine and anti-malarial dose recommendation in HIV-malaria co-treatment. Malar J. 2015;14:179.

39. Sambol NC, Yan L, Creek DJ, McCormack SA, Arinaitwe E, Bigira V, et al. Population pharmacokinetics of piperaquine in young Ugandan children treated with dihydroartemisinin-piperaquine for uncomplicated malaria. Clin Pharmacol Ther. 2015;98:87–95.

40. Huang L, Carey V, Lindsey JC, Marzan F, Gingrich D, Graham B, et al. Concomitant nevirapine impacts pharmacokinetic exposure to the antimalarial artemether-lumefantrine in African children. PLoS One. 2017;12:e0186589.

41. UNAIDS. In danger: UNAIDS global AIDS update 2022. Geneva: Joint United Nations Programme on HIV/AIDS; 2022.

42. WHO. Consolidated guidelines on HIV prevention, testing, treatment, service delivery and monitoring: recommendations for a public health approach. Geneva: World Health Organization; 2021.

43. Gasasira AF, Kamya MR, Ochong EO, Vora N, Achan J, Charlebois E, et al. Effect of trimethoprim-sulphamethoxazole on the risk of malaria in HIV-infected Ugandan children living in an area of widespread antifolate resistance. Malar J. 2010;9:177.

44. Kamya MR, Gasasira AF, Achan J, Mebrahtu T, Ruel T, Kekitiinwa A, et al. Effects of trimethoprim-sulfamethoxazole and insecticide-treated bednets on malaria among HIV-infected Ugandan children. AIDS. 2007;21:2059–66.

45. Gasasira AF, Kamya MR, Achan J, Mebrahtu T, Kalyango JN, Ruel T, et al. High risk of neutropenia in HIV-infected children following treatment with artesunate plus amodiaquine for uncomplicated malaria in Uganda. Clin Infect Dis. 2008;46:985–91.

46. Hewitt K, Steketee R, Mwapasa V, Whitworth J, French N. Interactions between HIV and malaria in non-pregnant adults: evidence and implications. AIDS. 2006;20:1993–2004.

47. Smart LR, Orgenes N, Mazigo HD, Minde M, Hokororo A, Shakir M, et al. Malaria and HIV among pediatric inpatients in two Tanzanian referral hospitals: a prospective study. Acta Trop. 2016;159:36–43.

48. Roberds A, Ferraro E, Luckhart S, Stewart VA. HIV-1 impact on malaria transmission: a complex and relevant global health concern. Front Cell Infect Microbiol. 2021;11:656938.

49. Ashley EA, Dhorda M, Fairhurst RM, Amaratunga C, Lim P, Suon S, et al. Spread of artemisinin resistance in Plasmodium falciparum malaria. N Engl J Med. 2014;371:411–23.

50. WHO. Strategy to respond to antimalarial drug resistance in Africa. Geneva: World Health Organization; 2022.

51. Byakika-Kibwika P, Lamorde M, Mayito J, Nabukeera L, Namakula R, Mayanja-Kizza H, et al. Significant pharmacokinetic interactions between artemether/lumefantrine and efavirenz or nevirapine in HIV-infected Ugandan adults. J Antimicrob Chemother. 2012;67:2213–21.

52. Byakika-Kibwika P, Lamorde M, Okaba-Kayom V, Mayanja-Kizza H, Katabira E, Hanpithakpong W, et al. Lopinavir/ritonavir significantly influences pharmacokinetic exposure of artemether/lumefantrine in HIV-infected Ugandan adults. J Antimicrob Chemother. 2012;67:1217–23.

53. German P, Parikh S, Lawrence J, Dorsey G, Rosenthal PJ, Havlir D, et al. Lopinavir/ritonavir affects pharmacokinetic exposure of artemether/lumefantrine in HIV-uninfected healthy volunteers. J Acquir Immune Defic Syndr. 2009;51:424–9.

54. Usman SO, Oreagba IA, Akinyede AA, Agbaje EO, Akinleye MO, Onwujuobi AG, et al. Effect of nevirapine, efavirenz and lopinavir/ritonavir on the therapeutic concentration and toxicity of lumefantrine in people living with HIV at Lagos University Teaching Hospital, Nigeria. J Pharmacol Sci. 2020;144:95–101.

55. Francis J, Barnes KI, Workman L, Kredo T, Vestergaard LS, Hoglund RM, et al. An individual participant data population pharmacokinetic meta-analysis of drug-drug interactions between lumefantrine and commonly used antiretroviral treatment. Antimicrob Agents Chemother. 2020;64:e02394–19.

56. Kawuma AN, Walimbwa SI, Pillai GC, Khoo S, Lamorde M, Wasmann RE, et al. Dolutegravir pharmacokinetics during co-administration with either artemether/lumefantrine or artesunate/amodiaquine. J Antimicrob Chemother. 2021;76:1269–72.

57. Walimbwa SI, Lamorde M, Waitt C, Kaboggoza J, Else L, Byakika-Kibwika P, et al. Drug Interactions between dolutegravir and artemether-lumefantrine or artesunate-amodiaquine. Antimicrob Agents Chemother. 2019;63:e01310–18.

58. Maganda BA, Minzi OM, Ngaimisi E, Kamuhabwa AA, Aklillu E. CYP2B6*6 genotype and high efavirenz plasma concentration but not nevirapine are associated with low lumefantrine plasma exposure and poor treatment response in HIV-malaria-coinfected patients. Pharmacogenomics J. 2016;16:88–95.

59. Arinaitwe E, Sandison TG, Wanzira H, Kakuru A, Homsy J, Kalamya J, et al. Artemether-lumefantrine versus dihydroartemisinin-piperaquine for falciparum malaria: a longitudinal, randomized trial in young Ugandan children. Clin Infect Dis. 2009;49:1629–37.

60. Van Geertruyden JP, Van Eijk E, Yosaatmadja F, Kasongo W, Mulenga M, D’Alessandro U, et al. The relationship of Plasmodium falciparum humeral immunity with HIV-1 immunosuppression and treatment efficacy in Zambia. Malar J. 2009;8:258.

61. Usman SO, Oreagba IA, Kadri MR, Adewumi OO, Akinyede A, Agbaje EO, et al. Evaluation of the effects of atazanavir-ritonavir on the pharmacokinetics of lumefantrine in patients living with HIV in Lagos University Teaching Hospital, South-Western Nigeria. Eur J Clin Pharmacol. 2021;77:1341–8.

62. Adegbola A, Abutaima R, Olagunju A, Ijarotimi O, Siccardi M, Owen A, et al. Effect of pregnancy on the pharmacokinetic interaction between efavirenz and lumefantrine in HIV-malaria coinfection. Antimicrob Agents Chemother. 2018;62:e01252–18.

63. Adegbola AJ, Rana A, Adeagbo BA, Bolarinwa RA, Olagunju AE, Siccardi M, et al. Influence of selected polymorphisms in disposition genes on lumefantrine pharmacokinetics when coadministered with efavirenz. Pharmacogenet Genomics. 2020;30:96–106.

64. Banda CG, Chaponda M, Mukaka M, Mulenga M, Hachizovu S, Kabuya JB, et al. Efficacy and safety of artemether-lumefantrine as treatment for Plasmodium falciparum uncomplicated malaria in adult patients on efavirenz-based antiretroviral therapy in Zambia: an open label non-randomized interventional trial. Malar J. 2019;18:180.

