## Supplementary material for "Efficacy of artemisinin-based combination therapy (ACT) in people living with HIV (PLHIV) diagnosed with uncomplicated *Plasmodium* falciparum malaria in Africa: A WWARN systematic review": Suppl_File_preprint: Suppl_File1_4-4-2025.docx

**Additional File 1. Search terms for the original search in MEDLINE, EMBASE, Web of Science (all Databases), Cochrane Central, WHO Global Index Medicus and Clinicaltrials.gov**

1	exp malaria/

2	exp Plasmodium/

3	malaria*.mp.

4	falciparum.mp.

5	1 or 2 or 3 or 4

6	exp Africa/

7	((Africa or Algeria or Angola or Benin or Botswana or "Burkina Faso" or Burundi or Cameroon or "Canary Islands" or "Cape Verde" or "Central African Republic" or Chad or Comoros or Congo or "Democratic Republic of Congo" or Djibouti or Egypt or "Equatorial Guinea" or Eritrea or Ethiopia or Gabon or Gambia or Ghana or Guinea or "Guinea Bissau" or "Ivory Coast" or "Cote d'Ivoire" or Jamahiriya or Kenya or Lesotho or Liberia or Libya or Madagascar or Malawi or Mali or Mauritania or Mauritius or Mayotte or Morocco or Mozambique or Namibia or Niger or Nigeria or Principe or Reunion or Rwanda or "Sao Tome" or Senegal or Seychelles or "Sierra Leone" or Somalia or "South Africa" or "St Helena" or Sudan or Swaziland or Tanzania or Togo or Tunisia or Uganda or "Western Sahara" or Zaire or Zambia or Zimbabwe or "Central* Africa*" or "West* Africa*" or "East* Africa*" or "North* Africa*" or "South* Africa*" or "sub Saharan Africa*" or "subSaharan Africa*") not ("guinea pig*" or "aspergillus niger")).mp. (358942)

8	6 or 7

9	artemisinin/ or artemisinin derivative/

10	artemether/

11	benflumetol/

12	artesunate plus pyronaridine/ or amodiaquine plus artesunate/ or artesunate plus mefloquine/ or artesunate plus chlorproguanil plus dapsone/

13	dihydroartemisinin plus piperaquine/ or dihydroartemisinin/ or dihydroartemisinin derivative/ (0)

14	piperaquine/

15	artemisinin*.mp.

16	Coartem.mp.

17	artemether.mp.

18	lumefantrin*.mp.

19	(artesunate and amodiaquine).mp.

20	(dihydroartemisinin and piperaquine).mp.

21	(artesunate and mefloquine).mp.

22	(artesunate and sulphadoxine and pyrimethamine).mp.

23	9 or 10 or 11 or 12 or 13 or 14 or 15 or 16 or 17 or 18 or 19 or 20 or 21 or 22

24	exp Human immunodeficiency virus 1 infection/ or exp Human immunodeficiency virus infection/ or exp acquired immune deficiency syndrome/ or exp Human immunodeficiency virus/ or immune deficiency/

25	"immunodeficiency syndrome*".mp.

26	"human immunodeficiency".mp.

27	"human t cell lymphotropic".mp.

28	HIV.mp.

29	"immunodeficiency virus*".mp.

30	"acquired immune deficiency".mp.

31	"immun* deficiency syndrome*".mp.

32	"acquired immunodeficienc*".mp.

33	aids.mp.

34	24 or 25 or 26 or 27 or 28 or 29 or 30 or 31 or 32 or 33

35	5 and 8 and 23 and 34

**Search Results**

| Ovid Medline | 115 |
| --- | --- |
| Ovid Embase | 308 |
| Ovid Global Health | 85 |
| Ebsco Cinahl | 8 |
| Scopus | 230 |
| Web of Science Core Collection | 117 |
| The Cochrane Library | 78 |
| Total | 941 |
