## Supplementary material for "Efficacy of artemisinin-based combination therapy (ACT) in people living with HIV (PLHIV) diagnosed with uncomplicated *Plasmodium* falciparum malaria in Africa: A WWARN systematic review": Suppl_File_preprint: Suppl_File2_4-4-2025.docx

**Additional File 2. Search terms for the subsequent searches in the WWARN Clinical Trial Library**

### Search Results

|  | 30/10/2020 search results | 01/07/2021 search results (2020 to present) | 28/04/2022 search results (2021 to current date only) |
| --- | --- | --- | --- |
| Ovid Medline | 1964 | 1433 | 1283 |
| Ovid Embase | 3331 | 1684 | 1455 |
| Web of Science (All Databases) | 4894 | 2524 | 2153 |
| Cochrane CENTRAL | 991 | 203 | 161 |
| WHO Global Index Medicus | 119 | 62 | 23 |
| Clinicaltrials.gov | 3574 | 3662 | 4024 [*not limited by date*] |
| Total | 14873 | 9568 | 9099 |
| Total after deduplication | 5841 | 1262 | 2435 |

### Search Strategies

**Database: Medline (Ovid MEDLINE® Epub Ahead of Print, In-Process & Other Non-Indexed Citations, Ovid MEDLINE® Daily and Ovid MEDLINE®) 1946 to present**

Search Strategy:

--------------------------------------------------------------------------------

1	exp Malaria/ (66741)

2	exp Plasmodium/ (47352)

3	malaria.ti,ab. (79322)

4	plasmodium.ti,ab. (49207)

5	1 or 2 or 3 or 4 (105499)

6	(AL or Amalar or Amatem or Amodiaquine or Ariplus or Arteether or Artefenomel or Artekin or artemether or Artemisinin or artemisone or artemotil or Artequick or Artequin or Arterolane or arterolane or Artesinin or artesun or artesunate or ASAQ or ASMQ or Atovaquone or avloclor or Azithromycin or Benzolol or Chloroquine or Chlorproguanil or cinchoine or cinchonidine or cipargamin or clindamycin or Coarsucam or Coartem or Co-arsucam or Co-artem or cotrimoxazole or co-trimoxazole or cycloguanil or dapsone or Dawaquin or Dihydroartemisinin or DHA or dihydroartimisininne or Doxycycline or DSM265 or Duo-Cotecxin or DuoCotecxin or eurartesim or Eurartesim or Fansidar or Fansimef or Faverid or Ferroquine or Fosmidomycin or Halfan or Halofantrine or hydroxychloroquine or Imatinib or ivermectin or KAE609 or KAF156 or Krintafel or Lariam or Lonart or lumefantrine or Malarone or Mefloquine or Mepron or Methylene Blue or MMV390048 or Monodox or naphthoquine or napthoquinone or nivaquine or OZ439 or Paludrine or piperaguine or piperaquine or Piperazine or pipraquine or primaquine or proguanil or Pyramax or pyramax or pyrimethamine or Pyronaridine or Qualaquin or quinidine or Quinimax or quinine or Resochin or Riamet or SB-252263 or SAR97276SP or SPAQ or SPAQ-CO or sulfadoxine or sulfamethoxypyridazine or sulphadoxine or sulphamethoxypyridazine or SYNRIAM or tafenoquine or tetracycline or TQ or Vibramycin or Vibramycin-D or WR238605).ti,ab. (436388)

7	hydroxychloroquine*.ti,ab. (5445)

8	hydroxy-chloroquine*.ti,ab. (48)

9	Vibramycin*.ti,ab. (191)

10	(P218 or SJ733 or ACT-451840 or "CDRI 9778" or M5717 or MMV253 or Atoguanil or AQ13 or Sevuparin or MMV048 or Ganaplacide).ti,ab. (52)

11	6 or 7 or 8 or 9 or 10 (436430)

12	5 and 11 (19779)

13	limit 12 to dt=20180701-20201231 (1964)

28/04/2022: limit 13 to yr="2021 -Current"

**Database: Embase 1974 to present**

Search Strategy:

--------------------------------------------------------------------------------

1	exp malaria/ (89113)

2	exp Plasmodium/ (60795)

3	malaria.ti,ab. (92634)

4	plasmodium.ti,ab. (56039)

5	1 or 2 or 3 or 4 (127875)

6	(AL or Amalar or Amatem or Amodiaquine or Ariplus or Arteether or Artefenomel or Artekin or artemether or Artemisinin or artemisone or artemotil or Artequick or Artequin or Arterolane or arterolane or Artesinin or artesun or artesunate or ASAQ or ASMQ or Atovaquone or avloclor or Azithromycin or Benzolol or Chloroquine or Chlorproguanil or cinchoine or cinchonidine or cipargamin or clindamycin or Coarsucam or Coartem or Co-arsucam or Co-artem or cotrimoxazole or co-trimoxazole or cycloguanil or dapsone or Dawaquin or Dihydroartemisinin or DHA or dihydroartimisininne or Doxycycline or DSM265 or Duo-Cotecxin or DuoCotecxin or eurartesim or Eurartesim or Fansidar or Fansimef or Faverid or Ferroquine or Fosmidomycin or Halfan or Halofantrine or hydroxychloroquine or Imatinib or ivermectin or KAE609 or KAF156 or Krintafel or Lariam or Lonart or lumefantrine or Malarone or Mefloquine or Mepron or Methylene Blue or MMV390048 or Monodox or naphthoquine or napthoquinone or nivaquine or OZ439 or Paludrine or piperaguine or piperaquine or Piperazine or pipraquine or primaquine or proguanil or Pyramax or pyramax or pyrimethamine or Pyronaridine or Qualaquin or quinidine or Quinimax or quinine or Resochin or Riamet or SB-252263 or SAR97276SP or SPAQ or SPAQ-CO or sulfadoxine or sulfamethoxypyridazine or sulphadoxine or sulphamethoxypyridazine or SYNRIAM or tafenoquine or tetracycline or TQ or Vibramycin or Vibramycin-D or WR238605).ti,ab. (876004)

7	hydroxychloroquine*.ti,ab. (9648)

8	hydroxy-chloroquine*.ti,ab. (99)

9	Vibramycin*.ti,ab. (156)

10	(P218 or SJ733 or ACT-451840 or "CDRI 9778" or M5717 or MMV253 or Atoguanil or AQ13 or Sevuparin or MMV048 or Ganaplacide).ti,ab. (110)

11	6 or 7 or 8 or 9 or 10 (876100)

12	5 and 11 (29202)

13	12 (29202)

14	limit 13 to yr="2018 -Current" (3331)

28/04/2022: limit 12 to yr="2021-Current"

**Web of Science All Databases:**

Web of Science Core Collection (1900-present)

Web of Science Core Collection: Citation Indexes

Science Citation Index Expanded (SCI-EXPANDED) --1900-present

Social Sciences Citation Index (SSCI) --1900-present

Arts & Humanities Citation Index (A&HCI) --1975-present

Conference Proceedings Citation Index- Science (CPCI-S) --1990-present

Conference Proceedings Citation Index- Social Science & Humanities (CPCI-SSH) --1990-present

Book Citation Index– Science (BKCI-S) --2005-present

Book Citation Index– Social Sciences & Humanities (BKCI-SSH) --2005-present

Emerging Sources Citation Index (ESCI) --2015-present

Web of Science Core Collection: Chemical Indexes

Current Chemical Reactions (CCR-EXPANDED) --1985-present

(Includes Institut National de la Propriete Industrielle structure data back to 1840)

Index Chemicus (IC) --1993-present

BIOSIS Citation Index (1969-present)

Current Contents Connect (1998-present)

Data Citation Index (1993-present)

Derwent Innovations Index (1993-present)

KCI-Korean Journal Database (1980-present)

MEDLINE® (1950-present)

Russian Science Citation Index (2005-present)

SciELO Citation Index (2002-present)

Zoological Record (1993-present)

Search Strategy:

--------------------------------------------------------------------------------

1. TOPIC: (malaria)
2. TOPIC: (plasmodium)
3. TOPIC: (AL or Amalar or Amatem or Amodiaquine or Ariplus or Arteether or Artefenomel or Artekin or artemether or Artemisinin or artemisone or artemotil or Artequick or Artequin or Arterolane or arterolane or Artesinin or artesun or artesunate or ASAQ or ASMQ or Atovaquone or avloclor or Azithromycin or Benzolol or Chloroquine or Chlorproguanil or cinchoine or cinchonidine or cipargamin or clindamycin or Coarsucam or Coartem or Co-arsucam or Co-artem or cotrimoxazole or co-trimoxazole or cycloguanil or dapsone or Dawaquin or Dihydroartemisinin or DHA or dihydroartimisininne or Doxycycline or DSM265 or Duo-Cotecxin or DuoCotecxin or eurartesim or Eurartesim or Fansidar or Fansimef or Faverid or Ferroquine or Fosmidomycin or Halfan or Halofantrine or hydroxychloroquine or Imatinib or ivermectin or KAE609 or KAF156 or Krintafel or Lariam or Lonart or lumefantrine or Malarone or Mefloquine or Mepron or Methylene Blue or MMV390048 or Monodox or naphthoquine or napthoquinone or nivaquine or OZ439 or Paludrine or piperaguine or piperaquine or Piperazine or pipraquine or primaquine or proguanil or Pyramax or pyramax or pyrimethamine or Pyronaridine or Qualaquin or quinidine or Quinimax or quinine or Resochin or Riamet or SB-252263 or SAR97276SP or SPAQ or SPAQ-CO or sulfadoxine or sulfamethoxypyridazine or sulphadoxine or sulphamethoxypyridazine or SYNRIAM or tafenoquine or tetracycline or TQ or Vibramycin or Vibramycin-D or WR238605)
4. TOPIC: (hydroxychloroquine* or hydroxy-chloroquine*)
5. TOPIC: (Vibramycin*)
6. TOPIC: (P218 or SJ733 or ACT-451840 or "CDRI 9778" or M5717 or MMV253 or Atoguanil or AQ13 or Sevuparin or MMV048 or Ganaplacide)
7. #2 OR #1
8. #6 OR #5 OR #4 OR #3
9. #8 AND #7
10. #8 AND #7 Refined by: PUBLICATION YEARS: ( 2020 OR 2019 OR 2018 )

28/04/2022: Refined by: **PUBLICATION YEARS:** ( 2021 OR 2022 )

**Cochrane Central Register of Controlled Trials**

**Issue 10 of 12, October 2020**

Search Strategy:

--------------------------------------------------------------------------------

#1	MeSH descriptor: [Malaria] explode all trees	2768

#2	MeSH descriptor: [Plasmodium] explode all trees	936

#3	malaria	6830

#4	plasmodium	3067

#5	#1 or #2 or #3 or #4	6947

#6	(AL or Amalar or Amatem or Amodiaquine or Ariplus or Arteether or Artefenomel or Artekin or artemether or Artemisinin or artemisone or artemotil or Artequick or Artequin or Arterolane or arterolane or Artesinin or artesun or artesunate or ASAQ or ASMQ or Atovaquone or avloclor or Azithromycin or Benzolol or Chloroquine or Chlorproguanil or cinchoine or cinchonidine or cipargamin or clindamycin or Coarsucam or Coartem or Co-arsucam or Co-artem or cotrimoxazole or co-trimoxazole or cycloguanil or dapsone or Dawaquin or Dihydroartemisinin or DHA or dihydroartimisininne or Doxycycline or DSM265 or Duo-Cotecxin or DuoCotecxin or eurartesim or Eurartesim or Fansidar or Fansimef or Faverid or Ferroquine or Fosmidomycin or Halfan or Halofantrine or hydroxychloroquine or Imatinib or ivermectin or KAE609 or KAF156 or Krintafel or Lariam or Lonart or lumefantrine or Malarone or Mefloquine or Mepron or Methylene Blue or MMV390048 or Monodox or naphthoquine or napthoquinone or nivaquine or OZ439 or Paludrine or piperaguine or piperaquine or Piperazine or pipraquine or primaquine or proguanil or Pyramax or pyramax or pyrimethamine or Pyronaridine or Qualaquin or quinidine or Quinimax or quinine or Resochin or Riamet or SB-252263 or SAR97276SP or SPAQ or SPAQ-CO or sulfadoxine or sulfamethoxypyridazine or sulphadoxine or sulphamethoxypyridazine or SYNRIAM or tafenoquine or tetracycline or TQ or Vibramycin or Vibramycin-D or WR238605)	114121

#7	hydroxychloroquine*	1465

#8	hydroxy-chloroquine*	13

#9	Vibramycin*	40

#10	P218 or SJ733 or ACT-451840 or "CDRI 9778" or M5717 or MMV253 or Atoguanil or AQ13 or Sevuparin or MMV048 or Ganaplacide	26

#11	#6 or #7 or #8 or #9 or #10	114140

#12	#5 and #11	4476

Custom Range: 01/07/2018 to 31/12/2020

28/04/2022: Year Custom Range: 2021 to 2022

**WHO Global Index Medicus**

[**https://pesquisa.bvsalud.org/gim/**](https://pesquisa.bvsalud.org/gim/)

Search Strategy:

--------------------------------------------------------------------------------

(tw:(malaria or plasmodium)) AND (tw:(AL or Amalar or Amatem or Amodiaquine or Ariplus or Arteether or Artefenomel or Artekin or artemether or Artemisinin or artemisone or artemotil or Artequick or Artequin or Arterolane or arterolane or Artesinin or artesun or artesunate or ASAQ or ASMQ or Atovaquone or avloclor or Azithromycin or Benzolol or Chloroquine or Chlorproguanil or cinchoine or cinchonidine or cipargamin or clindamycin or Coarsucam or Coartem or Co-arsucam or Co-artem or cotrimoxazole or co-trimoxazole or cycloguanil or dapsone or Dawaquin or Dihydroartemisinin or DHA or dihydroartimisininne or Doxycycline or DSM265 or Duo-Cotecxin or DuoCotecxin or eurartesim or Eurartesim or Fansidar or Fansimef or Faverid or Ferroquine or Fosmidomycin or Halfan or Halofantrine or hydroxychloroquine or Imatinib or ivermectin or KAE609 or KAF156 or Krintafel or Lariam or Lonart or lumefantrine or Malarone or Mefloquine or Mepron or Methylene Blue or MMV390048 or Monodox or naphthoquine or napthoquinone or nivaquine or OZ439 or Paludrine or piperaguine or piperaquine or Piperazine or pipraquine or primaquine or proguanil or Pyramax or pyramax or pyrimethamine or Pyronaridine or Qualaquin or quinidine or Quinimax or quinine or Resochin or Riamet or SB-252263 or SAR97276SP or SPAQ or SPAQ-CO or sulfadoxine or sulfamethoxypyridazine or sulphadoxine or sulphamethoxypyridazine or SYNRIAM or tafenoquine or tetracycline or TQ or Vibramycin or Vibramycin-D or WR238605 or hydroxychloroquine* or hydroxy-chloroquine* or Vibramycin* or P218 or SJ733 or ACT-451840 or "CDRI 9778" or M5717 or MMV253 or Atoguanil or AQ13 or Sevuparin or MMV048 or Ganaplacide))

Limited: 2018 – 2020

28/04/2022: Limited 2021- 2022

**Clinicaltrials.gov**

Search Strategy:

--------------------------------------------------------------------------------

Condition or disease: Malaria

Other terms (all searched individually):

AL or Amalar or Amatem or Amodiaquine or Ariplus or Arteether or Artefenomel or Artekin or artemether or Artemisinin or artemisone or artemotil or Artequick or Artequin or Arterolane or Artesinin or artesun or artesunate or ASAQ or ASMQ or Atovaquone or avloclor or Azithromycin or Benzolol or Chloroquine or Chlorproguanil or cinchoine or cinchonidine or cipargamin or clindamycin or Coarsucam or Coartem or Co-arsucam or Co-artem or cotrimoxazole or co-trimoxazole or cycloguanil or dapsone or Dawaquin or Dihydroartemisinin or DHA or dihydroartimisininne or Doxycycline or DSM265 or Duo-Cotecxin or DuoCotecxin or eurartesim or Eurartesim or Fansidar or Fansimef or Faverid or Ferroquine or Fosmidomycin or Halfan or Halofantrine or hydroxychloroquine or Imatinib or ivermectin or KAE609 or KAF156 or Krintafel or Lariam or Lonart or lumefantrine or Malarone or Mefloquine or Mepron or Methylene Blue or MMV390048 or Monodox or naphthoquine or napthoquinone or nivaquine or OZ439 or Paludrine or piperaguine or piperaquine or Piperazine or pipraquine or primaquine or proguanil or Pyramax or pyramax or pyrimethamine or Pyronaridine or Qualaquin or quinidine or Quinimax or quinine or Resochin or Riamet or SB-252263 or SAR97276SP or SPAQ or SPAQ-CO or sulfadoxine or sulfamethoxypyridazine or sulphadoxine or sulphamethoxypyridazine or SYNRIAM or tafenoquine or tetracycline or TQ or Vibramycin or Vibramycin-D or WR238605 or hydroxychloroquine or P218 or SJ733 or ACT-451840 or "CDRI 9778" or M5717 or MMV253 or Atoguanil or AQ13 or Sevuparin or MMV048 or Ganaplacide

No date limits possible
