## Supplementary material for "Efficacy of artemisinin-based combination therapy (ACT) in people living with HIV (PLHIV) diagnosed with uncomplicated *Plasmodium* falciparum malaria in Africa: A WWARN systematic review": Suppl_File_preprint: Suppl_File5_4-4-2025.docx

**Additional File 5. Inclusion criteria for enrolment of people living with HIV (PLHIV) in each study included in the review and their baseline CD4 measurements**

| Study ID | N: PLHIV enrolled | HIV-related inclusion criteria | CD4 count at enrolment |
| --- | --- | --- | --- |
| 1 | 19 | HIV-1 positive at 2-time points AND  NPV-based ART OR no ART treatment for >=4 weeks before enrolment | Not done |
| 2 | 269 | HIV-1 positive AND  >2 months on 200mg NVP BID or 600mg EFV daily OR No treatment | EFV-based ART: 298x10^6^ cells/L (9-694) median (IQR)  NVP-based ART: 354x10^6^ cells/L (19-1781) median (IQR)  No Treatment: 402x10^6^ cells/L (66-964) median (IQR) |
| 3 | 35 | No restriction | Not done |
| 4 | 12 | No restriction | Not done |
| 5 | 32 | HIV-exposed and currently breastfeeding OR HIV-confirmed mother/child | Not done |
| 6 | 44 | HIV-exposed and currently breastfeeding OR HIV-confirmed mother/child | Not done |
| 7 | N/A ^1^ | HIV-confirmed mother/child | Not done |
| 8 | 166 | HIV-infected AND  ART-naïve and eligible for ART treatment OR already on ART treatment | ART-naïve (study1): 16% (2-44) median (IQR)  ART-naïve (study2): 21% (2-61) median (IQR)  ART-experienced (study1): 30% (8-51) median (IQR)  ART-experienced (study2): NA |
| 9 | 170 | HIV-confirmed infection AND  ART-naïve OR eligible for initiation of ART OR receiving standard first-line ART > 12 months AND viral load <400 copies/mL in last 6 months | No Treatment: 14-16% (2-44) median (IQR)  First Line ART: 30-31% (8-51) median (IQR) |
| 10 | 36 | HIV-1-confirmed infection AND  ART-naïve OR on EFV-based ART for >=4 weeks before enrolment | No treatment: 301 cells/µL (256-346) mean (95%CI)  EFV-based ART: 382 cells/µL (310-454) mean (95%CI) |
| 11 | 118 | No restriction | Not done |
| 12 | 320 | Pre-study HIV status unknown | HIV1-infected: 256 cells/µL (170-406) median (IQR)  HIV uninfected: NA |
| 13 | 10 | HIV1-confirmed infection AND  receiving ATV/r-based ART treatment AND CD4 >=200 cells/mm^3^ | Not done |
| 14 | 69 | HIV-confirmed infection AND  >=2 weeks EFV-based ART | Pregnant women: 469.4 cells/µL (210.5) mean (SD)  Non-pregnant women: 406 cells/µL (308.5) mean (SD) |
| 15 | 9 | HIV-confirmed infection with 2 assays AND  initiation of EFV-based ART >=10 days before enrolment | Not done |
| 16 | 30 | HIV-confirmed infection AND  stabilized on current ART treatment | Not done |
| 17 | 152 | HIV-confirmed infection AND  >=24 weeks EFV-based ART | EFV-based ART: 376 cells/µL (248-511) median (IQR) |
| 18 | 221 | HIV-confirmed infection AND  receiving NPV-based ART or EFV-based ART | EFV-based ART: 256 cells/mm^3^ (140-360) median (IQR)  NVP-based ART: 390 cells/mm^3^ (237-500) median (IQR) |
| 19 | 68 | HIV-confirmed infection AND  receiving NVP-based ART | Not done |

^1^ Not reported, only total malaria episodes (n=38) among PLHIV reported and analyzed

Study ID: 19 studies included in the review and labelled as in Table 1 of the main manuscript.

ART: antiretroviral therapy; ATV/r: atazanavir-ritonavir; CI: confidence interval; EFV: efavirenz; IQR: interquartile range; NVP: nevirapine; SD: standard deviation
