## Supplementary material for "Efficacy of artemisinin-based combination therapy (ACT) in people living with HIV (PLHIV) diagnosed with uncomplicated *Plasmodium* falciparum malaria in Africa: A WWARN systematic review": Suppl_File_preprint: Suppl_File7_4-4-2025.docx

**Additional File 7. Summary of the reported day 7 lumefantrine concentrations (ng/mL)**

| Study ID | Publication reference | HIV treatment | N | Specific Group | Median | Geometric Mean | Mean | SEM | IQR | Range | 95%CI | 90%CI | N below 280 ng/mL (%) |
| --- | --- | --- | --- | --- | --- | --- | --- | --- | --- | --- | --- | --- | --- |
| 2 | (1) | EFV | N/A |  | 300.4 |  |  |  | 220.8- 343.1 |  |  |  |  |
| 9 | (2) | EFV | 25 |  | 97 |  |  |  | 61-124 |  |  |  |  |
| 10 | (3) | EFV+TS | 22 |  |  |  | 260 |  | 163-357 ^5^ |  | 200-320 |  |  |
| 11 | (4) | EFV | 48 |  | 111 |  |  |  | 63-192 |  |  |  |  |
| 9 | (2) | LPV/r | 65 |  | 926 |  |  |  | 473-1910 |  |  |  |  |
| 11 | (4) | LPV/r | 70 |  | 1140 |  |  |  | 515-2220 |  |  |  |  |
|  |  | HIV-uninfected group | 186 |  | 340 |  |  |  | 257-531 |  |  |  |  |
| 1 | (5) | HIV-uninfected group | 20 |  | 323 |  |  |  | 164- 404 ^4^ | 53.9-779 |  |  |  |
| 2 | (1) | None | N/A |  | 970 |  |  |  | 562.1-1729 |  |  |  |  |
| 1 | (5) | NVP | 15 |  | 697 |  |  |  | 374-1100 ^4^ | 155-2250 |  |  |  |
| 2 | (1) | NVP | N/A |  | 1125 |  |  |  | 638.8-1913 |  |  |  |  |
| 9 | (2) | NVP | 67 |  | 388 |  |  |  | 164-563 |  |  |  |  |
| 11 | (4) | NVP | 62 |  | 426 |  |  |  | 282-733 |  |  |  |  |
| 10 | (3) | TS | 22 |  |  |  | 640 |  | 479-801 ^5^ |  | 540-740 |  |  |
| 13 | (6) | ATV/r | 10 |  |  |  | 3847 | 894 |  |  | 2096-5598 ^5^ |  |  |
|  |  | HIV-uninfected group | 10 |  |  |  | 1375 | 266 |  |  | 854-1896 ^5^ |  |  |
| 14 | (7) | EFV | 27 | Pregnant women | 279 |  |  |  | 120-610 |  |  |  |  |
|  |  | EFV | 25 | Non-pregnant women | 212 |  |  |  | 133-400 |  |  |  |  |
| 14 | (8) | EFV | 11 | NR1I3 1089 TT* | 146.1 |  |  |  |  |  | 83.6-313.2 |  | 70 |
|  |  | EFV | 42 | NR1I3 1089 TC and CC* | 271.1 |  |  |  |  |  | 144.7-479.6 |  | 53 |
|  |  | EFV | 20 | CYP2B6 516 GG* | 289.4 |  |  |  |  |  | 148.1-569.9 |  | 47 |
|  |  | EFV | 33 | CYP2B6 516 TT and GT* | 211.7 |  |  |  |  |  | 123.1-352 |  | 59 |
| 15 | (9) | HIV-uninfected group | 30 | Pregnant women |  | 409 |  |  |  |  |  | 231-617 |  |
|  |  | EFV | 9 | Pregnant women |  | 160 |  |  |  |  |  | 134-309 |  |
| 16 | (10) | None | 10 |  | 290 |  |  |  | 275-304 |  |  |  |  |
|  |  | NVP | 10 |  | 369 |  |  |  | 337-502 |  |  |  |  |
|  |  | EFV | 10 |  | 239 |  |  |  | 213-243 |  |  |  |  |
|  |  | LPV/r | 10 |  | 1331 |  |  |  | 1015-1903 |  |  |  |  |
| 17 | (11) | EFV | 85 ^1^ |  | 240 |  |  |  | 143.2-370 |  |  |  |  |
| 19 | (12) | HIV-uninfected group | 94 |  | 1455 ^2^ |  |  |  | 545-2280 |  |  |  |  |
|  |  | NVP | 68 |  | 1878 ^3^ |  |  |  | 1095-2840 |  |  |  |  |

*patients grouped according to cytochromes P450: CYP2B6 516 GG, CYP2B6 516 GT and CYP2B6 516 TT genotypes and nuclear receptors NR1I3 152c-1089: TC and TT alleles

^1^ an additional 36 patients were below the lower limit of quantification (<50 ng/mL)

^2^ reported as 2.75 (IQR 1.03-4.31) µM, a molecular weight of 528.939 g/mol was used in the calculation; there were also 5 participants with extremely low readings not included in the calculation

^3^ reported as 3.55 (IQR 2.07-5.37) µM, a molecular weight of 528.939 g/mol was used in the calculation

^4^ estimated using the methods described by Hozo, *et al*. (13)

^5^ calculated using <http://vassarstats.net/median_range.htm>

ATV/r: atazanavir-ritonavir; CI: confidence interval (90 or 95%); EFV: efavirenz; IQR: interquartile range; LPV/r: lopinavir-ritonavir; NVP: nevirapine; N/A: information not available; SEM: standard error of the mean; TS: trimethoprim-sulfamethoxazole preventive treatment
