## Supplementary figures and images for "Efficacy of artemisinin-based combination therapy (ACT) in people living with HIV (PLHIV) diagnosed with uncomplicated *Plasmodium* falciparum malaria in Africa: A WWARN systematic review"

### Suppl_File6_4-4-2025.tiff

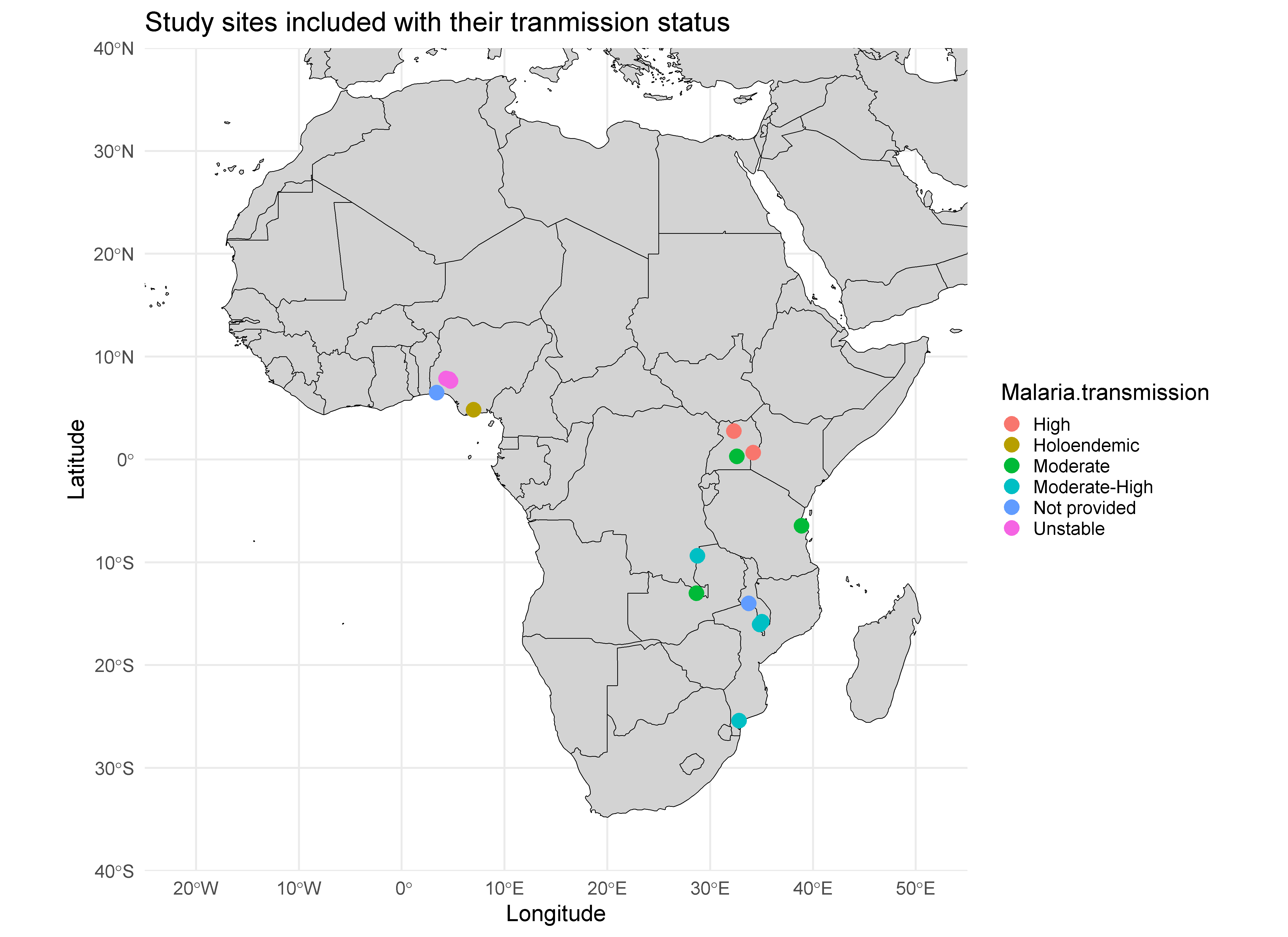
